# Increased Frequency of Clonal Hematopoiesis of Indeterminate Potential in Bloom Syndrome Probands and Carriers

**DOI:** 10.1101/2024.02.02.24302163

**Authors:** Isabella Lin, Angela Wei, Tsumugi A Gebo, PC Boutros, Maeve Flanagan, Nicole Kucine, C Cunniff, VA Arboleda, VY Chang

**Affiliations:** Department of Human Genetics, David Geffen School of Medicine, UCLA, Los Angeles, CA, USA; Department of Pathology and Laboratory Medicine, David Geffen School of Medicine, UCLA, Los Angeles, CA, USA; Department of Computational Medicine, David Geffen School of Medicine, UCLA; Interdepartmental BioInformatics Program, UCLA; Department of Urology, University of California Los Angeles, Los Angeles, CA; Jonsson Comprehensive Cancer Center, UCLA, Los Angeles, CA; Institute for Precision Health, University of California Los Angeles, Los Angeles, CA; Department of Pediatrics, Weill Cornell Medicine, New York, NY; Molecular Biology Institute, University of California, Los Angeles, CA; Division of Pediatric Hematology/ Oncology, UCLA, Los Angeles, CA; Children’s Discovery and Innovation Institute, UCLA, Los Angeles, CA

## Abstract

**Background:** Bloom Syndrome (BSyn) is an autosomal recessive disorder caused by biallelic germline variants in *BLM,* which functions to maintain genomic stability. BSyn patients have poor growth, immune defects, insulin resistance, and a significantly increased risk of malignancies, most commonly hematologic. The malignancy risk in carriers of pathogenic variants in *BLM* (*BLM* variant carriers) remains understudied. Clonal hematopoiesis of indeterminate potential (CHIP) is defined by presence of somatic mutations in leukemia-related genes in blood of individuals without leukemia and is associated with increased risk of leukemia. We hypothesize that somatic mutations driving clonal expansion may be an underlying mechanism leading to increased cancer risk in BSyn patients and *BLM* variant carriers.

**Methods:** To determine whether *de novo* or somatic variation is increased in BSyn patients or carriers, we performed and analyzed exome sequencing on BSyn and control trios.

**Results:** We discovered that both BSyn patients and carriers had increased numbers of low-frequency, putative somatic variants in CHIP genes compared to controls. Furthermore, BLM variant carriers had increased numbers of somatic variants in DNA methylation genes compared to controls. There was no statistical difference in the numbers of *de novo* variants in BSyn probands compared to control probands.

**Conclusion:** Our findings of increased CHIP in BSyn probands and carriers suggest that one or two germline pathogenic variants in *BLM* could be sufficient to increase the risk of clonal hematopoiesis. These findings warrant further studies in larger cohorts to determine the significance of CHIP as a potential biomarker of aging, cancer, cardiovascular disease, morbidity and mortality.

## INTRODUCTION

Bloom syndrome (BSyn, OMIM #210900) is a rare autosomal recessive disorder characterized by prenatal and postnatal growth restriction, sun sensitivity, insulin resistance, mild immune deficiency, and increased risk of early-onset malignancy^1–3^. Predominantly, BSyn cases are caused by homozygous or compound heterozygous variants in the *BLM* gene (OMIM# 604611), with over 70 different pathogenic variants identified, including several founder variants in Ashkenazi, Slavic, Spanish and Portuguese populations^4,5^.

The BLM protein, a 3’->5’ ATP-dependent RecQ DNA helicase, plays crucial roles in genome maintenance, including the stabilization of replication forks^3^, suppression of DNA crossovers during meiosis^6^, disentanglement of under-replicated DNA strands during metaphase, and management of complex DNA structures such as G complexes^7^. Notably, cells from individuals with BSyn exhibit an excess number of sister-chromatid exchanges (SCE), historically considered the diagnostic test for BSyn.^8–11^ However, increased SCE has not proven predictive of cancer development and there are currently no evidence-based cancer surveillance guidelines and biomarkers for early cancer detection^8,12^.

The heightened incidence of early-onset cancer in BSyn patients is thought to be driven by genomic instability resulting from biallelic germline variants in *BLM*^13–16^. For example, a recent BSyn patient with T-cell anaplastic large-cell lymphoma showed an excess of inter- and intra-chromosomal structural rearrangements, 150 and 1,293 respectively, in tumor cells^17^.

The Bloom Syndrome Registry reported that 53% of participants had developed cancer, with hematologic malignancies being the most common^16^. The most common solid tumors included colon, breast, and oropharyngeal cancers. The well-established cancer risk in BSyn individuals contrasts with the conflicting literature surrounding cancer risk in carriers of pathogenic variants in *BLM* (*BLM* variant carriers)^5,18–23^. A study conducted in New York and Israeli Ashkenazi Jews identified an increased risk of colorectal cancer in *BLM* variant carriers, with an odds ratio of 2.34^24^, and an Australian study in a high-risk breast cancer cohort identified a heterozygous, deleterious, protein truncating variant in *BLM* that co-segregated with cancer in a family^19^. A Netherlands cohort of early-onset colorectal cancer reported an increased frequency of heterozygous loss-of-function *BLM* variants^18^. Moreover, *in vitro* and *in vivo* experiments found that heterozygous *BLM* mutations increased susceptibility to mesothelioma^25^.

Clonal hematopoiesis of indeterminate potential (CHIP) is characterized by somatic mutations in leukemia-related “CHIP” genes detected in the blood of individuals without apparent hematologic malignancy^26,27^. The definition of CHIP has been a subject of ongoing discussion, ranging from a benign precursor state for hematologic neoplasms^28^, to the presence of a clonally expanded hematopoietic stem cell with a mutation with leukemogenic potential^29^. CHIP significantly elevates hematologic malignancy risk, with an annual increased risk of leukemia ranging from 0.5% to 1.0%^26^. Furthermore, CHIP has been implicated in adverse outcomes, such as cardiovascular disease, inflammation, and reduced age-adjusted life expectancy^30–32^.

The prevalence of CHIP varies with age, with up to 10% of patients aged 60-70 showing evidence of CHIP, while only 1% of those under the age of 50 seem to be affected. This age dependent pattern is attributed to the cumulative effects of mutations and telomere shortening in haematopoietic stem cells and multipotent progenitors over time, resulting in selective fitness and a profound decrease in clonal diversity^26,32–34^.

Studies suggest increased risk of CHIP in individuals harboring germline mutations in DNA repair and telomere maintenance genes^33,35,36^. In a study on heterozygous carriers of *POT1*, a gene involved in telomerase-dependent telomere elongation, predisposition to clonal hematopoiesis was observed in an autosomal dominant pattern of inheritance. This increased risk demonstrated age-dependent penetrance, with the hypothesis that longer telomere length allowed clones to sustain clonality^37^. Patients with short telomere syndrome have also been found to have an increased incidence of CHIP, although there is a wide spectrum of mutation types and genes in which these mutations occur^38,39^. This suggests that germline genetics can create a “permissive” environment for clonal evolution^38^, leading to clonal selection in hematopoietic cells.

Furthermore, in a recent large-scale study of UK Biobank, Memorial Sloan Kettering IMPACT, and The Cancer Genome Atlas cohorts, *BLM* was identified as one of 18 clonal hematopoiesis genes also associated with hematopoietic malignancy in the heterozygous state^40^.

Although clonal cytogenetic abnormalities in Bloom Syndrome fibroblast cell lines hint at a neoplastic process, no comprehensive genomic analyses have assessed overall somatic mutational load or CHIP as potential predictive biomarkers for hematologic malignancy^41,42^. We hypothesize that 1 or 2 germline *BLM* variants may heighten CHIP risk, potentially at earlier ages, correlating with increased malignancy risk identified in BSyn individuals and mildly elevated risk in *BLM* variant carriers. This study used exome sequencing to investigate the correlation between germline *BLM* variants, germline mutational load, and the risk of developing CHIP, potentially at earlier ages. To determine whether *de novo* or somatic variation occurs at rates in BSyn patients and *BLM* variant carriers different from control populations, we conducted exome analysis and variant annotation for these cohorts.

No significant differences in *de novo* mutation were identified in BSyn probands compared to controls; however, both BSyn probands and *BLM* variant carriers exhibited an increased incidence of low frequency, putatively somatic variants in CHIP and DNAm genes compared to sex- and age-matched controls. This study sheds new light on the interplay between genetic predispositions and somatic variation, and highlights the need for additional studies to further evaluate the mechanisms and potential clinical implications for patients with one or two *BLM* variants.

## METHODS

### Study Participants and Ethical Approval

All study participants provided informed consent under a protocol for the Bloom Syndrome Registry approved by the Weill Cornell Medical College Institutional Review Board, and a material transfer agreement was obtained.

### Exome Sequencing

We performed exome sequencing with the Nextera DNA Flex Pre-Enrichment Library Prep and the Roche NimbleGen exome capture kit following standard protocols. Libraries were indexed, multiplexed and sequenced on a 2x150 Illumina NovaSeq S1 flowcell at the UCLA Technology Center for Genomics and Bioinformatics.

Age- and sex-matched control trios were obtained from the publicly available dbGAP study phs001272.v1.p1 deposited by the Broad Institute Center for Mendelian Genomics. Control trios harbored undiagnosed disease without cancer phenotypes **(Table S1)** and samples were processed with the Illumina Nextera Exome Kit and sequenced on an Illumina HiSeq.

### Data Processing, Variant Calling, and Annotation

All FASTQ files underwent unified quality control, mapping and variant-calling based on GATK best-practices pipeline ^43,44^ (**Supplemental Methods, Figure S1A and S1B**).

Given the reliable identification of high-confidence somatic SNV/indel calls at positions with sufficient sequencing coverage >10X, variant calls were filtered to maintain read depth (DP)>10 over the alternate allele.

Family structure and variants were confirmed with *BLM* gene variant visualization using *Integrative Genomics Viewer v2.9.4* and variant annotation using *VarSeq 2.3.0*.

### Variant Filtering

Variants were initially filtered for DP and analyzed for coverage across regions of interest. Subsequent filtering for genotype quality (GQ) and a quality score of “PASS” were included (**Figure S1B, Supplemental Methods**).

### Variant Allele Frequency (VAF) Analysis

*De novo* variants were subsetted based on standard somatic and germline variant allele frequencies (VAF). VAF, representing the percentage of sequencing reads matching a specific DNA variant^45^, was used as a surrogate measure of allele proportion. A VAF<0.3 indicated acquired somatic variants, while VAF≥0.3 indicated likely germline or *de novo* variants^46–49^.

Given the range of VAF cutoffs in literature, analyses were also conducted using a more stringent VAF cutoff of 0.25, aligning with estimates of somatic singleton mutations in the range 0.1 ≤ VAF ≤ 0.25 from previous studies^50^.

## RESULTS

### Trio exome analysis for BSyn and control trios

We performed exome sequencing on 29 peripheral blood DNA samples obtained from the Bloom Syndrome Registry. The cohort consisted of 10 BSyn probands and their biological parents who are obligate carriers of these pathogenic variants in *BLM* (**Table 1**). Among the BSyn probands, there were equal numbers of male (n=5) and female (n=5), with ages ranging from 10 months to 36 years of age at the time of sample collection. Half (n=5) of the BSyn probands had a history of developing at least one type of cancer before or after the sample was collected (**Table 1**).

The pathogenic *BLM* variants observed in BSyn trios spanned amino acid 25 to 1243 of the *BLM* gene, with most variants clustered in the DEAH Helicase and RecQ Helicase C-terminal domains (**Figure 1A**). Family structure and *BLM* variant confirmation were ensured using *Integrative Genomics Viewer v2.9.4* (**Figure 1B**) and detailed variant annotation (**Table S2**).

**Figure 1:**
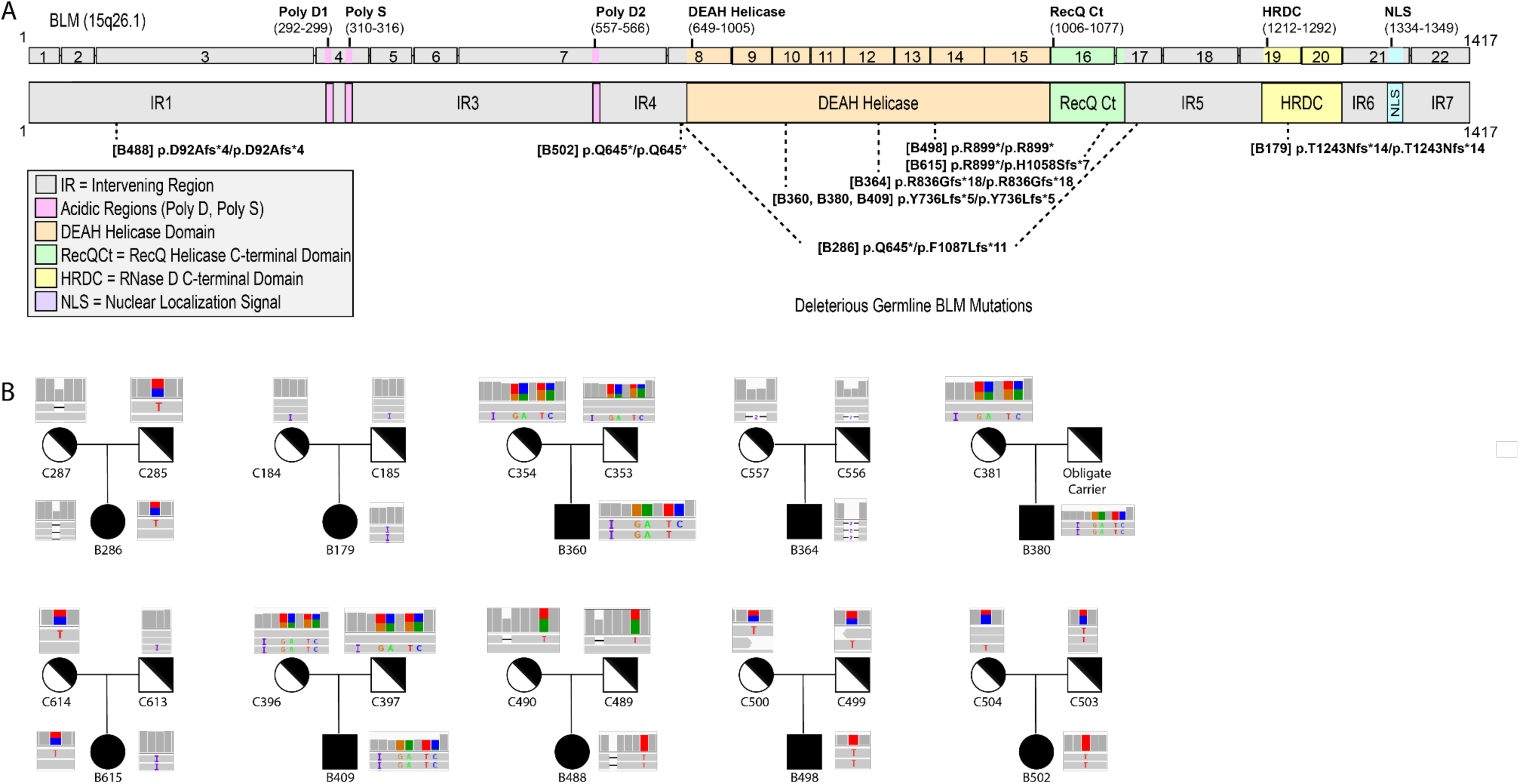
Genomic analysis of *BLM* Mutations in Bloom Syndrome Patients and Carriers. (**A**) Schematic representation of the *BLM* transcript (ENST00000355112.8) and protein (GenBank: BLM; NM000057.4; GRCh38), its functional domains (solid lines above transcript), and mutations (dotted lines below transcript) causing Bloom Syndrome. Variants listed correspond to BSyn probands in this study and are tagged with patient identifiers (B#, see Table 1). The location of deleterious biallelic variants are shown with one dotted line, while compound heterozygous variants are shown with two dotted lines. (**B**) Variants in each BSyn proband (B#) and BSyn carrier (C#) in our cohort were verified in *Integrative Genomic Viewer v.2.9.4*. Each *IGV* screenshot shows coverage at the *BLM* variant at the top and the first two to three sequencing reads below with reference bases in grey and genetic variants in color. Histograms represent the coverage around the Bsyn variants at that site. Each trio relationship is depicted.

No significant difference in mean total reads was identified between BSyn and control trio samples (**Figure S1C**, **Table S3**, t-test, *p*-value=0.268). After mapping to exome targets, BSyn trio samples had a mean of 113.1x coverage compared to controls with 106.5x coverage (**Figure S1D, Table S3**, t-test, *p*-value=0.018). This coverage consistency extended across all chromosomes (**Figure S1E**).

Samples were categorized into four key groups: BSyn proband samples (n=10) designated as “affected”, *BLM* variant carrier samples as “carrier” (n=19), and control proband (n=19) and control parents (n=38) as “unaffected.” In our somatic variant analysis, we separately considered control parent and control children as age-matched controls to assess the incidence of CHIP.

### BSyn probands and *BLM* variant carriers harbor somatic mutations in CHIP genes

We investigated the occurrence of low frequency, putative somatic variants (VAF < 0.3) in CHIP genes among BSyn probands and *BLM* variant carriers, revealing a median of 2, contrasting with control cohorts where no somatic CHIP gene variants were detected (median = 0) (Kruskal Wallis, *p*-value=1.50E-06 to 6.37E-03) **(Figure 2A**, **Table 2, Table S5)**. Applying a more stringent VAF cutoff of ≤ 0.25, based on differing cutoffs for somatic variants in literature, still identified more variants in CHIP genes in BSyn probands and *BLM* variant carriers compared to control cohorts, and notably significant comparisons between *BLM* variant carriers and control cohorts (Kruskal Wallis, *p*-value=8.82E-06 to 7.37E-03) (**Table S5** highlighted in pink).

**Figure 2:**
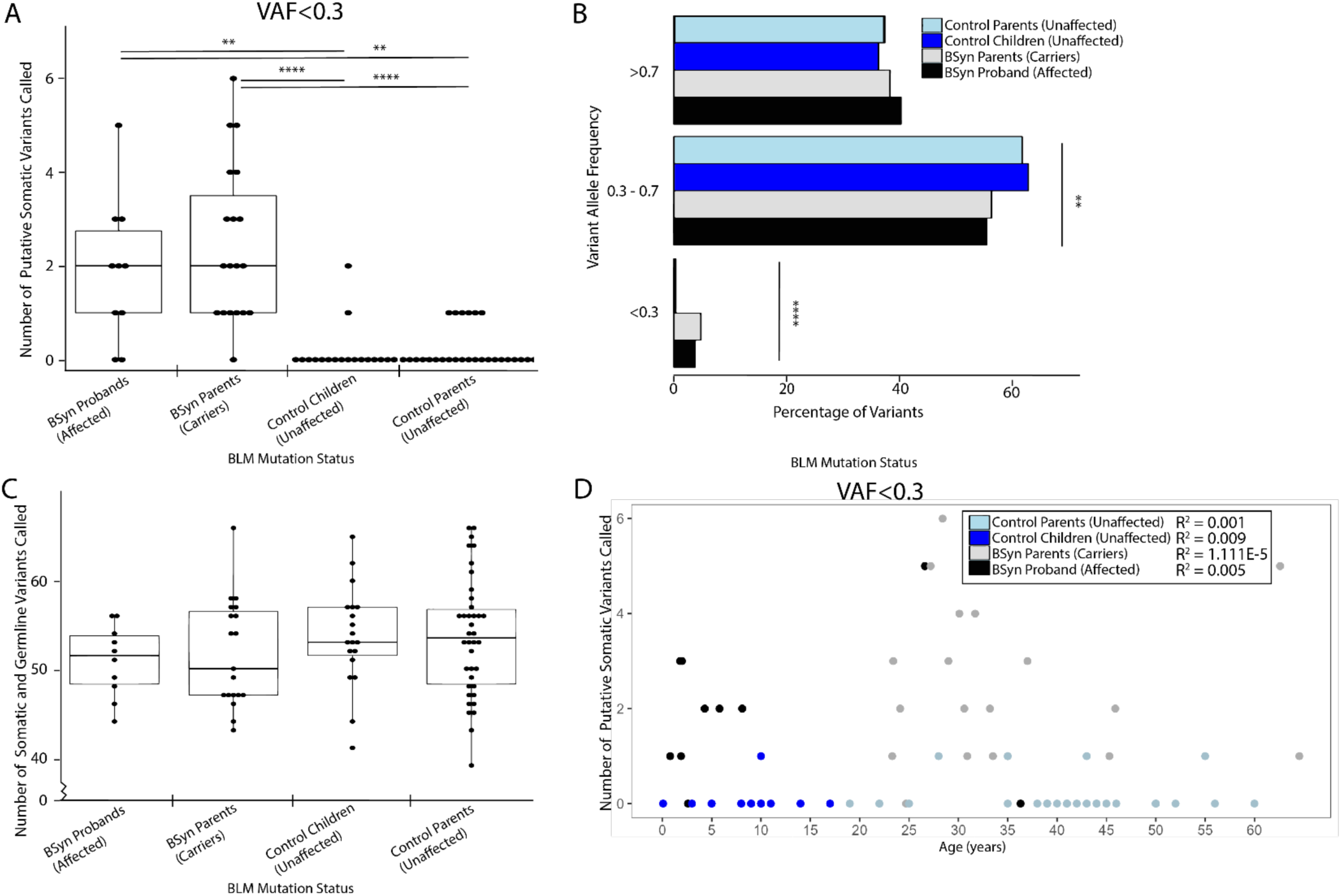
Comparative Analysis of Somatic CHIP Gene Variants Identified Significantly More in BSyn Probands and Carriers. We examined variants in CHIP genes in exome sequenced samples from four cohorts: BSyn probands (n=10, black), BSyn carrier parents (n=19, grey), control children (n=19, light blue) and control parents (n=38, dark blue). (**A**) Using VarSeq variant calling, we identified the number of putative somatic CHIP variants using a variant allele frequency (VAF) cutoff < 0.3 (Table 2). (**B**) CHIP gene variants were subsetted based on variant allele frequency (VAF) grouping, and the mean proportions are shown (Table S5). (**C**) The total number of germline and somatic CHIP gene variants are shown (Table 2). (**D**) We examined the effect of age on the number of putative somatic variants in CHIP genes that were called. Linear regression analysis was performed separately for each cohort and the R^2^ values indicate goodness of fit for each model: R^2^ = 0.005 (BSyn proband), R^2^ = 1.111E-05 (carrier), R^2^ = 0.009 (control child), R^2^ = 0.001 (control parent). These values suggest a weak linear relationship between age and the frequency of somatic CHIP variants in our cohorts. ns or no stars denote *p*-value>.05, * denote *p*-value≤.05, ** denote *p*-value<.01, *** denote *p*-value<.001, **** denote *p*-value<.0001

We explored multiple read depth cut-offs for loci in CHIP genes (**Supplemental Methods**). While DP>0 (**Figure S2A**) and DP>10 (**Figure S2B**) actually identified more loci in control samples than BSyn samples, DP>50 showed no significant difference in pairwise coverage comparison for total loci covered in CHIP genes (**Figure S2C**). A heatmap plotting all loci called for each sample at DP>0 confirmed nearly complete overlap across all samples (**Figure S2D**).

We further categorized variants in CHIP genes into putative somatic or germline based on VAF (**Figure 2B, Supplemental methods**). Consistently, significant differences were observed across all likely somatic variant comparisons between BSyn groups (model mean = 3.70 - 4.80%) and control groups (model mean = 0.30%) (*p*-value=1.41E-06 to 1.60E-03) (**Table S6**). No significant differences were found in the mean proportion of germline and somatic variants between BSyn probands and *BLM* variant carriers (*p*-value=0.447), nor between control probands and control parents (*p*-value=0.991) (**Figure 2B**).

Our analysis identified no significant correlations between mean somatic and germline variants in CHIP genes and the putative somatic subset (**Figure S3A**). Across the four sample groups, we identified no significant difference between mean somatic and germline number of variants in CHIP genes (**Figure 2C**, **Table 2**), type of variant (**Figure S3B**, Refseq Genes 110, NCBI), pathogenicity (**Figure S3C**, ClinVar 2023-01-05, NCBI), or CHIP genes to which these variants mapped (**Figure S3D**, Refseq Genes 110, NCBI). One-way ANOVA with random family effect confirmed that total CHIP gene variants followed a normal distribution (**Figure S4A**). No significant differences were observed in the type of variant (**Figure S4B**), variant pathogenicity (**Figure S4C**), or CHIP gene distribution (**Figure S4D**).

We also explored the influence of age on the number of putative somatic variants in CHIP genes in each cohort (**Figure 2D**). Linear regression analysis identified very weak linear relationships between age and the frequency of putative somatic CHIP variants at VAF<0.3 in our cohorts (R^2^ = 1.111E-05 to 0.009).

The increased presence of low frequency, putative somatic variants in CHIP genes for BSyn probands and *BLM* variant carriers is not attributable to differential coverage at CHIP genes but is likely due to increased somatic variant load in key cancer-related genes such as CHIP genes.

### BSyn probands and *BLM* variant carriers harbor somatic variants in DNA methylation genes

Epigenetic alterations have been linked to aging and cancer, and recent findings have found an accelerated aging, DNA methylation episignature in patients and mice with biallelic variants in *BLM* ^51–54^. Furthermore, loss of a different RecQ helicase protein, WRN, results in accelerated rates of DNA damage and increased DNA methylation aging in patients with Werner Syndrome who exhibit many clinical signs of accelerated aging^55,56^. To explore the impact of *BLM* mutations on mutational load in DNA methylation (DNAm) genes, we conducted a comprehensive analysis (**Methods, Supplemental methods**)^57,58^.

We identified significantly more low frequency, putatively somatic DNAm gene variants (VAF < 0.3) in BSyn probands and *BLM* variant carriers compared to unaffected control cohorts **(Figure 3A)**. *BLM* variant cohorts exhibited a median of 1 somatic DNAm gene variant, compared to a median of 0 in BSyn proband, control children and control parent cohorts. Statistically, these differences were significant in comparisons between *BLM* variant carrier and control children cohorts (Kruskal Wallis, *p*-value=7.941E-04) and *BLM* variant carrier and control parent cohorts (Kruskal Wallis, *p*-value=8.815E-04) (**Figure 3A**). These somatic mutations are listed in **Table S8**.

**Figure 3:**
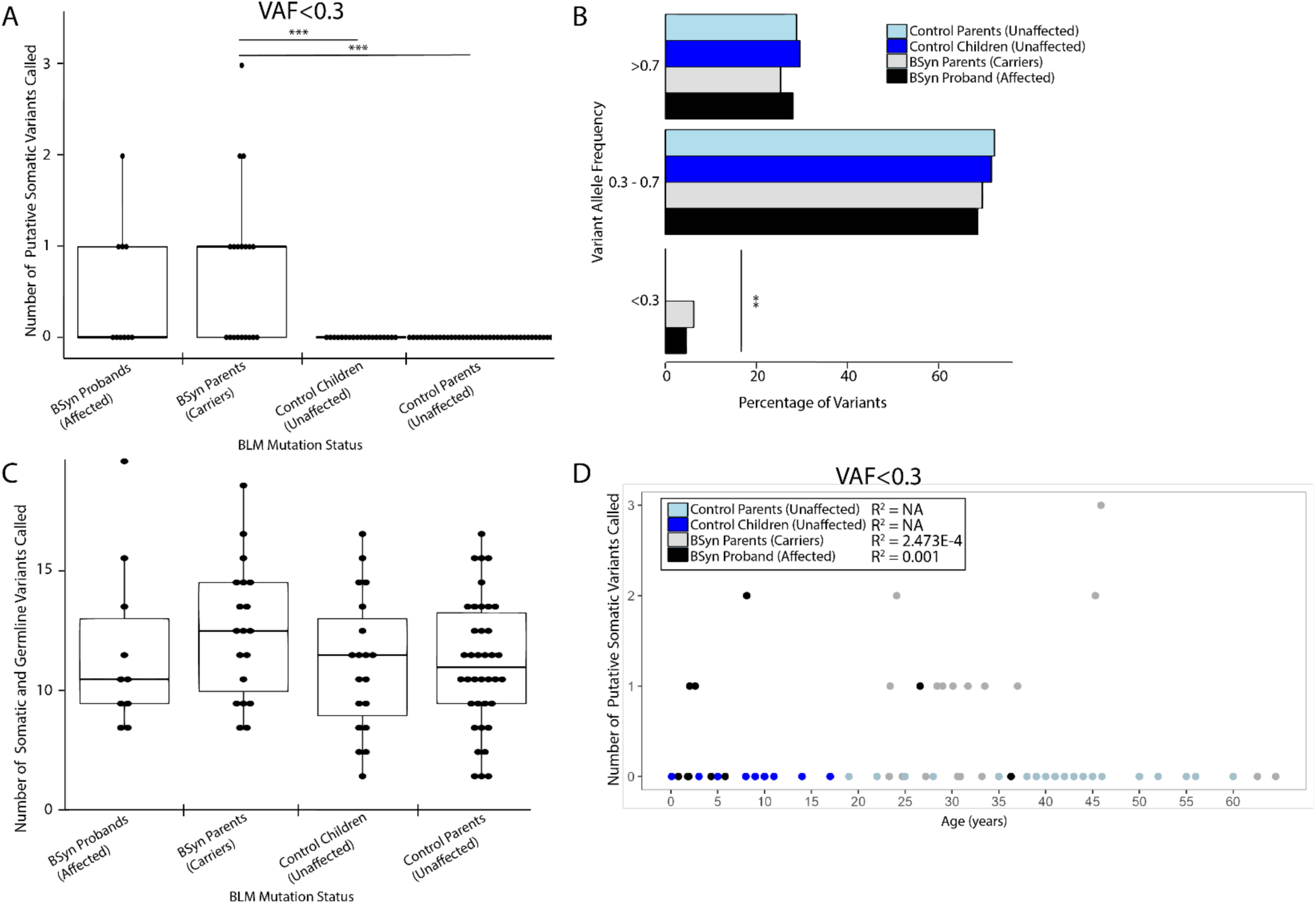
Comparative Analysis of Somatic DNA Methylation Gene Variants Identified Significantly More in BSyn Carriers Compared to Controls. We examined variants in DNAm genes in exome sequenced samples from four cohorts: BSyn probands (n=10, black), BSyn carrier parents (n=19, grey), control children (n=19, light blue) and control parents (n=38, dark blue). (**A**) Using VarSeq variant calling, we identified the number of putative somatic DNAm variants using a variant allele frequency (VAF) cutoff < 0.3 (Table S6). (**B**) DNAm gene variants were subsetted based on variant allele frequency (VAF) grouping, and the mean proportions are shown (Table S7). (**C**) The total number of germline and somatic DNAm gene variants are shown (Table S6). (**D**) We examined the effect of age on the number of putative somatic variants in DNAm genes that were called. Linear regression analysis was performed separately for each cohort and the R^2^ values indicate goodness of fit for each model: R^2^ = 0.001 (BSyn proband), R^2^ = 2.473E-04 (carrier), R^2^ = NA (control child), R^2^ = NA (control parent). These values suggest a weak to no linear relationship between age and the frequency of somatic CHIP variants in our cohorts. ns or no stars denote *p*-value>.05, * denote *p*-value≤.05, ** denote *p*-value<.01, *** denote *p*-value<.001, **** denote *p*-value<.0001

Applying a more stringent VAF cutoff of ≤ 0.25 still identified more variants in DNAm genes in BLM variant carriers compared to control cohorts, with significant differences between BLM variant carriers and the age-matched control cohort (Kruskal Wallis, *p*-value=0.044) (**Table S8** highlighted in pink).

We assessed multiple read depth cut-offs for loci in DNAm genes. While DP>0 (**Figure S5A**) and DP>10 (**Figure S5B**) identified more loci covered in control samples than BSyn samples, DP>50 showed no significant difference in pairwise coverage comparison for total loci in DNAm genes (**Figure S5C**). Loci coverage analysis identified almost complete overlap (**Figure S5D**). The DNAm gene list overlaps with the CHIP gene list as depicted (**Figure S6A, Table S4)**. Total variant analysis in DNAm genes was further explored, categorizing variants into putative somatic or germline based on VAF (**Figure 3B**). We identified significant differences in the mean proportions of variants, with higher proportions of somatic DNAm gene variants in BSyn proband (4.3%+/-2.3%) and *BLM* variant carrier (5.7%+/-1.9%) cohorts compared to control parent (0.0%+/-0.0%) cohorts (**Figure 3B, Table S9**). Significant differences were noted between *BLM* variant carriers and control children (mean proportion, *p*-value=2.944E-03), and *BLM* variant carriers and control parents (mean proportion, *p*-value=2.944E-03).

No significant differences were found in the mean number of total variants in DNAm genes using one way ANOVA with random family effect (**Figure 3C, Figure S6B**). Similarly, in our mean proportion analysis, no significant differences were observed based on the types of variants (**Figure S6C**) or DNAm gene (**Figure S6D**) in which the variant occurred. Additionally, the effect of age on the frequency of putative somatic variants in DNAm genes was assessed across cohorts (**Figure 3D**). Linear regression analysis identified very weak to no linear relationships between age and the frequency of putative somatic DNAm variants at VAF<0.3 in our cohorts (R2 = 0.001 to NA). These findings suggest that the increased presence of low frequency, putatively somatic DNAm gene variants in BSyn probands and *BLM* variant carriers is likely due to presence of the BLM variant and not age or family-effects.

### BSyn is not associated with increased *de novo* mutation rates

Finally, we assessed whether BLM mutations have differential effects on germline or somatic mutation rates, which are driven by different molecular mechanisms. One proband (B380) was excluded from this analysis as sequencing from only one parent was available. High quality coding variants in probands of each trio (n=9, Figure S7A) that were not inherited from either parent were identified, representing *de novo* variants (DNVs, VAF≥0.3) and newly arising somatic variants (VAF<0.3) (**Figure 4A**). No significant difference was found between BSyn and control cohorts (**Table S10, Table S11**, t-test, *p*-value=0.124).

**Figure 4:**
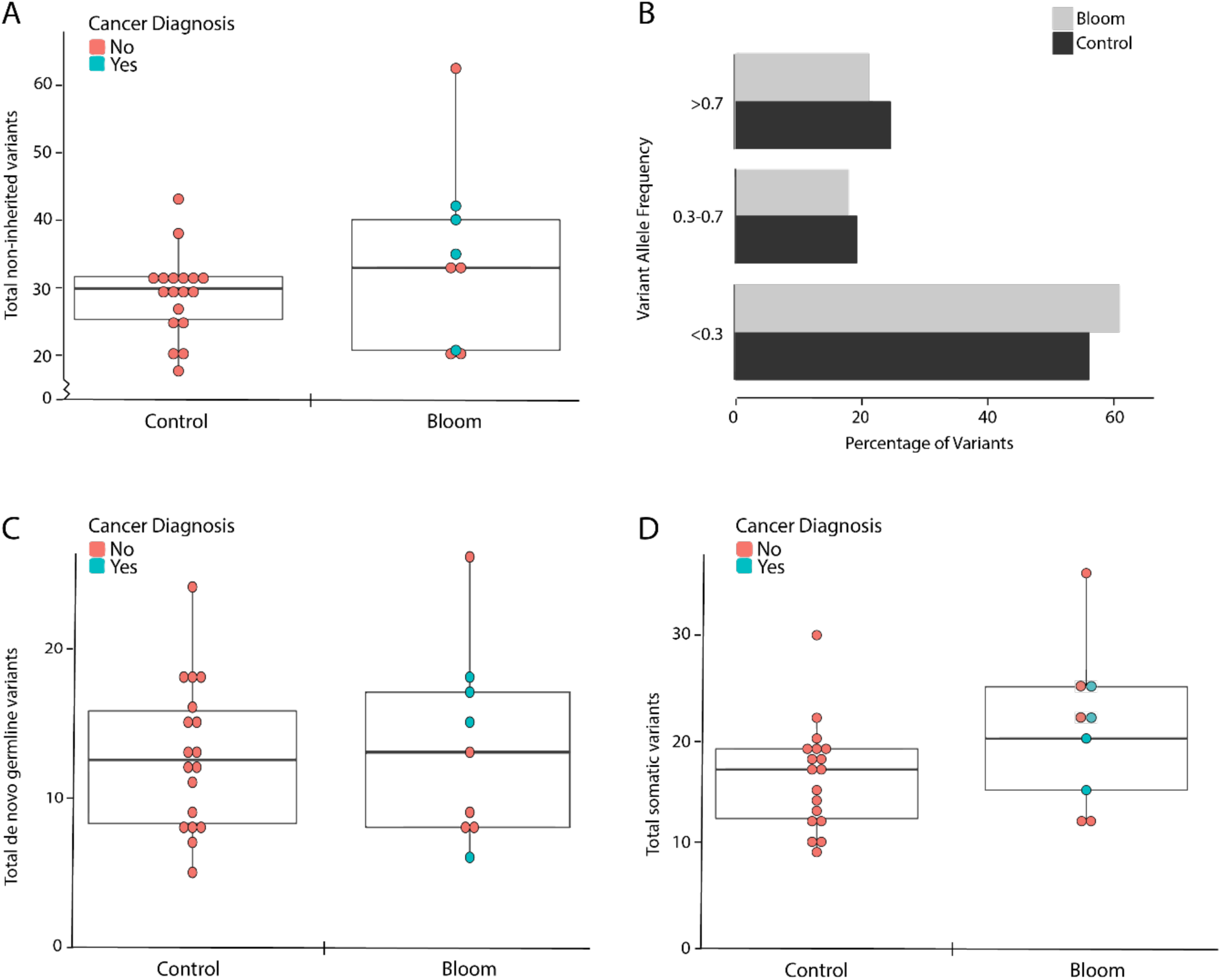
De Novo and Exome-Wide Somatic Variant Rates in BSyn and Controls. (**A**) We examined variants not inherited from either parent in our trio cohorts using VarSeq Trio Exome variant calling. This encompasses both germline *de novo* and newly arising somatic variants in BSyn probands (n=9) and age- and sex-matched control (n=18) (Table S10, Table S11). T-test was conducted to compare BSyn and control numbers of variants (t-test, *p*-value=0.187). (**B**) Non-inherited variants in BSyn probands and age- and sex-matched controls were subsetted based on variant allele frequency (VAF) grouping. The mean proportions are shown (Table S10, Table S11). (**C**) The number of germline *de novo* variants (VAF≥0.3) for each sample are shown (Table S11). T-test was conducted to compare BSyn and control numbers of variants (t-test, *p*-value=0.628). (**D**) The number of somatic variants (VAF<0.3) for each sample are shown (Table S9). T-test was conducted to compare BSyn and control numbers of variants (t-test, *p*-value=0.083). Pink indicates no cancer diagnosis, green indicates history of cancer. ns denotes *p*-value>0.05, implying no statistically significant difference between BSyn probands and controls.

Mean proportions of coding variants were assessed based on VAF categories, revealing no significant differences (**Figure 4B, Table S11**). The number and type of coding DNVs (VAF≥0.3) showed no significant difference between BSyn probands and control children (**Figure 4C, Figure S7B, Table S10**, t-test, p-value=0.628), aligning with reported literature values^59^. Similarly, putative somatic variants (VAF<0.3) showed no significant differences between BSyn probands and control children (**Figure 4D, Table S10**, t-test, *p*-value=0.083). However, the mean number of somatic variants for BSyn probands was 20.78, compared to a mean number of 16.33 in control children, suggesting a potential trend that a larger sample size would clarify.

We examined whether the *de novo* mutation rate was influenced by parental age at conception. An inverse relationship (slope=-0.961) between paternal age at conception and number of DNVs in the BSyn probands was observed, however this correlation was not robust(R^2^=0.355) (**Figure S7C**). This likely stems from a small sample size and a narrow range of paternal age at proband conception (19-32 years of age). No significant difference in DNVs was identified between BSyn probands with or without a cancer diagnosis. No significant trend was observed between the number of DNVs in BSyn probands with maternal age at conception (R^2^=0.294, data not shown) or with BSyn proband age at collection (R^2^=0.039) (**Figure S7D**).

## DISCUSSION

This study addresses the impact of pathogenic *BLM* variants on the incidence of *de novo* variants and somatic variants. Our findings reveal an increased frequency of low frequency, putatively somatic variants in CHIP genes (**Figure 2A**) and DNAm genes (**Figure 3A**) in BSyn probands and *BLM* variant carriers, compared to sex- and age-matched controls. These variants are predominantly represented by synonymous variants in BSyn probands and splice variants in *BLM* variant carriers (**Figure S3B**). In contrast to prior studies on clonal hematopoiesis, which identified predicted-pathogenic somatic variants predominantly in *DNMT3A*, *ASXL1*, and *TET2*, we identified mainly synonymous and benign splice variants primarily in *NOTCH1* and *CUX1*26,60,61.

The absence of significant differences in mean somatic and germline variants between BSyn probands and *BLM* variant carriers raises questions about whether BLM mutations alone drive these mutations. Other contributing factors, such as environmental exposures or other genetic modifiers, may influence the observed somatic mutation patterns. It is essential to acknowledge the study’s limitation of a small sample size and the effect on statistical significance. For example, although the mean number of exome-wide somatic variants for BSyn probands was higher than in control children, it did not attain statistical significance.

The findings of increased low-frequency putative somatic variants in CHIP genes aligns with the established roles of CHIP genes in cancer predisposition. This suggests a potential link between *BLM* mutations and an increased somatic mutation load in key cancer-related genes. An oral presentation at the American Society of Hematology 2023 Meeting further support this, identifying *BLM as* one of 18 clonal hematopoiesis genes associated with hematopoietic malignancy in the heterozygous state^40^, consistent with our findings of increased low-frequency putative somatic variants in *BLM* carriers.

We hypothesize that the increased number of putative somatic variants identified may result from reduced function of BLM causing genomic instability leading to an increased propensity for somatic mutations. These identified variants, largely synonymous and benign splice variants, may be a consequence of cell fitness selection, as they are less likely to be functional and more neutral to cell fitness and selection bias. Functional (deleterious) mutations would likely be selected against even in cells with deleterious *BLM* variants, while variants that promote growth and maintenance pathways may experience clonal expansion within the bone marrow. This expansion could contribute to a stable and functional hematopoietic system despite the underlying genetic alterations. Furthermore, cellular mechanisms involved in maintaining genomic stability, such as alternative DNA repair pathways or enhanced surveillance against accumulation of deleterious genetic alterations, could act as protective factors, potentially compensating for the deficiency caused by BLM mutations.

Our findings underscore the potential elevated risk for malignancies in BLM variant carriers, as evidenced by 63% (12/19) of *BLM* variant carriers exhibiting 2 or more somatic CHIP variants, a significant association compared to controls. However, the complex landscape of conflicting evidence on increased cancer risk in BLM variant carriers necessitates further investigation^20,23,62–65^. Deep-amplicon sequencing is recommended to verify and validate our findings of increased incidence of somatic variants observed through exome sequencing.

This study further explores the increased risk of CHIP in patients with germline mutations in genes involved in DNA repair and telomere maintenance genes. The *BLM* gene is a DNA helicase involved in replication fork stabilization and the suppression of homologous recombination, which could influence genomic stability, potentially contributing to the observed increase of somatic variants in CHIP-associated genes. This is consistent with existing evidence linking germline mutations in DNA repair and telomere maintenance genes and an increased risk of clonal hematopoiesis^33,35,36,38,39^.

Epigenome dysregulation is a hallmark of many diseases, including hematologic malignancies^26,61,66,67^. DNAm genes are known regulators of the dynamic epigenome^68^. We identified significantly increased numbers of somatic variants in DNAm genes in BSyn probands and *BLM* variant carriers. This prompts consideration of the potential impact of BLM mutations on accelerated aging and methylation changes as observed recently by Lee et al.^54^, similar to findings in patients with mutations in *WRN* ^69^. Unfortunately, insufficient DNA was available from each sample to perform DNAm assays. Our findings advocate for future studies and suggest that individuals with BLM mutations may benefit from sequencing-based screenings, which include many genes involved in DNAm processes^26,61,66,67^, for early detection of hematological malignancies.

Contrary to our expectations, we did not observe a significant difference in *de novo* mutation rates between BSyn probands and control cohorts. However, the increased mean number of somatic variants for BSyn probands implies that a larger sample size may provide clarity on *de novo* mutation rates in this population.

The study’s limitations include the rarity of Bloom Syndrome, resulting in a small sample size that constrains our ability to detect significant differences in *de novo* mutation rates and assess the potential impact of cancer therapy on the prevalence of clonal hematopoiesis. Although cancer treatments conceivably shape the landscape of clonal hematopoiesis^70^, our study did not find significant correlations between prior cancer diagnosis, including treatments administered before sample collection, and the number of somatic variants in BSyn patients (**Figure 4D**). An interesting follow-up would be to examine temporal samples, enabling monitoring of age-related expansions of potential somatic mutations and clarification of whether these represent true clonal somatic mosaicism. Such analyses could enhance statistical power in evaluating variant load in BSyn patients, and distinguishing those with and without cancer diagnoses.

The use of two different exome enrichment methods is another limitation, mitigated through joint computational processing. Additionally, the use of exome sequencing poses challenges in detecting ultra-low-frequency clones associated with CHIP variants, and the nature of exome sequencing with enrichment may lead to biases in coverage and detection of somatic variants. Our findings point to the potential utility of targeted assay for CHIP variants to address these ultra-low-frequency variants.

Finally, the association between true CHIP and cancer risk, as observed in various studies, may be influenced by factors such as smoking^26,61,71^. Some studies indicate a strong link between CHIP and a history of smoking, which may be a confounding effect on association studies. Although smoking history is unknown in our BSyn cohort, it is reasonable to assume that the BSyn probands did not have significant pack-year history given their ages.

The complex interplay between BLM mutations and the mutational landscape, revealing associations with somatic mutations in cancer-related genes and DNA methylation alterations, underscores the need for further investigation. Our findings contribute to the growing literature on the heightened somatic mutation rate and increased cancer risk in carriers for genes important in maintaining genomic integrity, such as *ATM* and a subset of Fanconi Anemia-related genes^72–74^. These findings may pave the way for early biomarkers in cancer detection and general health assessment in rare disease patients and carriers. Larger-scale studies with BSyn cohorts are imperative to unravel the mechanisms underpinning *BLM* loss-of-function variants and their contribution to CHIP and cancer risk.

### Public Data Sets

Control Exome data was obtained from Study ID: phs000178.v11.p8.c1, submitted by the Center for Mendelian Genomics [CMG] - The Broad Institute Joint Center for Mendelian Genomics. The Exomes used are: SRA ID SRS2136666, SRS2813808, SRS2136486, SRS2140039, SRS2140061, SRS2136721, SRS2203482, SRS2202906, SRS2202907, SRS2130875, SRS2136628, SRS2130876, SRS2197363, SRS2197826, SRS2197795, SRS2140305, SRS2137393, SRS2137389, SRS2200570, SRS2200550, SRS2200596, SRS2195820, SRS2195786, SRS2195798, SRS2200588, SRS2200627, SRS2200615, SRS2205811, SRS2195821, SRS2195834, SRS2136679, SRS2136619, SRS2136617, SRS2136629, SRS2136659, SRS2136613, SRS2203316, SRS2202950, SRS2202953, SRS2203490, SRS2203471, SRS2203336, SRS2200551, SRS2200573, SRS2200609, SRS2200624, SRS2200562, SRS2200610, SRS2200576, SRS2200561, SRS2288808, SRS2288810, SRS2288816, SRS2200626, SRS2200613, SRS2288805, SRS2200605

## Data Availability

All data produced in the present study are available upon reasonable request to the authors

## Acknowledgements

We thank the UCLA Technology for Genomics and Bioinformatics for their sequencing expertise, and Dr. Jeff Gornbein from the UCLA Statistics Department for his expertise and aid in statistical analyses. We would like to acknowledge the families and patients who participated in this study and donated their time and efforts to make this possible.

## Contribution Statement

VC, VA, IL and AW designed and conceptualized the study, analyzed the generated data, and wrote the paper. PB and TAG performed mapping and variant calling of WES data and IL performed VarSeq trio exome analysis. MF, NK, CC coordinated sample collection and DNA extraction. All authors contributed to the final editing of the manuscript.

## Table and Supplemental Table Legends

**Table 1: Bloom Syndrome Patient and *BLM* variant Carrier Demographics**

Bloom Syndrome patient (B#, n=10) and *BLM* variant carrier (C#, n=19) parents included in our studies are identified with their patient IDs. Individual sex, age range, family grouping, *BLM* mutation and prior history of cancer are also shown.

**Table 2: Statistical Calculations for CHIP Gene Variant Analysis**

One way (mixed) analysis of variance (ANOVA) model with a random family effect to allow for non-independence between cohorts was conducted for the total number of CHIP gene variants. The mean and median number of variants for each sample group, as well as the standard deviation (SD), pooled standard error of mean (SEM), and degrees of freedom (df) are shown. For each comparison, the adjusted p values are listed.

The non-parametric Kruskal-Wallis method was conducted for the somatic CHIP gene variants across the four sample groupings. The mean and median number of variants for each sample group, as well as the standard deviation (SD) are shown. For each comparison, the adjusted p values are listed.

**Table S1: Control Children and Parent Demographics**

Control children (n=19) and parents (n=38) included in our studies are identified with their sample IDs and SRA accession numbers from dbGaP. Individual sex, age range at sample collection, family grouping, genetic disorder and sequencing information are shown for samples where sample information was provided.

**Table S2: Identification of deleterious BLM Mutations in Samples**

Using VarSeq, we identified all deleterious variants in *BLM* in our BSyn proband (n=10), *BLM* variant carrier (n=19), and control cohorts (n=57). We identified no deleterious variants in any of the control individuals, as expected, and confirmed expected heterozygous deleterious variants in *BLM* in *BLM* variant carriers. For BSyn probands, we confirmed the expected homozygous or compound heterozygous deleterious variants in *BLM*.

**Table S3: Sequencing Coverage**

For each sample, we show sequencing coverage, mapping statistics, and exome depth of coverage. These were generated using the Broad Institute Picard Metrics.

**Table S4: Clonal Hematopoiesis of Indeterminate Potential (CHIP) Genes and DNA methylation (DNAm) Genes**

CHIP genes were compiled from Schenz et al.^27^ and Hagiwara et al.^34^ DNAm genes were compiled from Rasmussen & Helin ^57^ and Ginno et al.^58^ These genes were used for our analysis of CHIP genes and DNAm gene variants respectively.

**Table S5: High Quality Variants Detected at VAF < 0.3 in CHIP Genes**

VarSeq was used to detect high quality variants. All non-reference variants with VAF < 0.3 in CHIP genes are listed here with corresponding read depth, gene name and exon, effect of variant, sample ID and age range. Variants with VAF < 0.25 are highlighted in light pink.

**Table S6: Proportion Calculations for CHIP Gene Variants**

A multinomial logistic model with a random person effect was conducted to analyze mean proportions of CHIP gene variants in each cohort based on variant allele frequency (VAF) groupings. The observed and model mean proportions, as well as standard errors are listed for each sample group. For each comparison, the difference, standard error of difference, and adjusted p values are listed.

**Table S7: Statistical Calculations for DNAm Gene Variant Analysis**

One way (mixed) analysis of variance (ANOVA) model with a random family effect to allow for non-independence between cohorts was conducted for the total number of DNAm gene variants. The mean and median number of variants for each sample group, as well as the standard deviation (SD), pooled standard error of mean (SEM), and degrees of freedom (df) are shown. For each comparison, the adjusted p values are listed.

The non-parametric Kruskal-Wallis method was conducted for the somatic DNAm gene variants across the four sample groupings. The mean and median number of variants for each sample group, as well as the standard deviation (SD) are shown. For each comparison, the adjusted p values are listed.

**Table S8: High Quality Variants Detected at VAF < 0.3 in DNAm Genes**

VarSeq was used to detect high quality variants. All non-reference variants with VAF < 0.3 in DNAm genes are listed here with corresponding read depth, gene name and exon, effect of variant, sample ID and age range. Variants with VAF < 0.25 are highlighted in light pink.

**Table S9: Proportion Calculations for DNAm Gene Variants**

A multinomial logistic model with a random person effect was conducted to analyze mean proportions of DNAm gene variants in each cohort based on variant allele frequency (VAF) groupings. The observed and model mean proportions, as well as standard errors are listed for each sample group. For each comparison, the difference, standard error of difference, and adjusted p values are listed.

**Table S10: Statistical Calculations for Trio Exome Variant Analysis**

Pairwise t-test was conducted at each variant filtering step (Figure S1A) to compare numbers of variants between Bloom Syndrome probands (n=9) and control children (n=18) at each filtering step.

**Table S11: Trio Variant Annotation and Breakdown**

Bloom syndrome and control trio samples were processed as per Varseq best practices Trio Exome pipeline. Sample ID, age ranges and sex are shown with the number of variants remaining for each individual proband after each set of filtering (see Methods and Figure S1B) are listed. Variants werefurther categorized into mode of inheritance - transmitted, *de novo*, or MIE (mendel inheritance error). *De novo* variants are further broken down by VAF (variant allele frequency).

## REFERENCES

1. German J, Archibald R, Bloom D. CHROMOSOMAL BREAKAGE IN A RARE AND PROBABLY GENETICALLY DETERMINED SYNDROME OF MAN. Science 1965;148(3669):506–507.

2. Bloom D. Congenital telangiectatic erythema resembling lupus erythematosus in dwarfs; probably a syndrome entity. AMA Am J Dis Child 1954;88(6):754–758.

3. Cunniff C, Bassetti JA, Ellis NA. Bloom’s Syndrome: Clinical Spectrum, Molecular Pathogenesis, and Cancer Predisposition. Mol Syndromol 2017;8(1):4–23.

4. Ellis NA, Ciocci S, Proytcheva M, Lennon D, Groden J, German J. The Ashkenazic Jewish Bloom syndrome mutation blmAsh is present in non-Jewish Americans of Spanish ancestry. Am J Hum Genet 1998;63(6):1685–1693.

5. Sokolenko AP, Iyevleva AG, Preobrazhenskaya EV, et al. High prevalence and breast cancer predisposing role of the BLM c.1642 C>T (Q548X) mutation in Russia. Int J Cancer 2012;130(12):2867–2873.

6. Wu L, Hickson ID. The Bloom’s syndrome helicase suppresses crossing over during homologous recombination. Nature 2003;426(6968):870–874.

7. Manthei KA, Keck JL. The BLM dissolvasome in DNA replication and repair. Cell Mol Life Sci 2013;70(21):4067–4084.

8. Chaganti RS, Schonberg S, German J. A manyfold increase in sister chromatid exchanges in Bloom’s syndrome lymphocytes. Proc Natl Acad Sci U S A 1974;71(11):4508–4512.

9. Onoda F, Seki M, Miyajima A, Enomoto T. Elevation of sister chromatid exchange in Saccharomyces cerevisiae sgs1 disruptants and the relevance of the disruptants as a system to evaluate mutations in Bloom’s syndrome gene. Mutat Res 2000;459(3):203–209.

10. McDaniel LD, Schultz RA. Elevated sister chromatid exchange phenotype of Bloom syndrome cells is complemented by human chromosome 15. Proc Natl Acad Sci U S A 1992;89(17):7968–7972.

11. Wang W, Seki M, Narita Y, et al. Functional relation among RecQ family helicases RecQL1, RecQL5, and BLM in cell growth and sister chromatid exchange formation. Mol Cell Biol 2003;23(10):3527–3535.

12. Dicken CH, Dewald G, Gordon H. Sister chromatid exchanges in Bloom’s syndrome. Arch Dermatol 1978;114(5):755–760.

13. Hodson C, Low JKK, van Twest S, et al. Mechanism of Bloom syndrome complex assembly required for double Holliday junction dissolution and genome stability. Proc Natl Acad Sci U S A;119(6.):

14. German J, Crippa LP, Bloom D. Bloom’s syndrome. III. Analysis of the chromosome aberration characteristic of this disorder. Chromosoma 1974;48(4):361–366.

15. German J. Bloom’s syndrome. XX. The first 100 cancers. Cancer Genet Cytogenet 1997;93(1):100–106.

16. Sugrañes TA, Flanagan M, Thomas C, Chang VY, Walsh M, Cunniff C. Age of first cancer diagnosis and survival in Bloom syndrome. Genet Med 2022;24(7):1476–1484.

17. Ye X, Maglione PJ, Wehr C, et al. Genomic characterization of lymphomas in patients with inborn errors of immunity. Blood Adv [Epub ahead of print].

18. de Voer RM, Hahn M-M, Mensenkamp AR, et al. Deleterious Germline BLM Mutations and the Risk for Early-onset Colorectal Cancer. Sci Rep 2015;514060.

19. Thompson ER, Doyle MA, Ryland GL, et al. Exome sequencing identifies rare deleterious mutations in DNA repair genes FANCC and BLM as potential breast cancer susceptibility alleles. PLoS Genet 2012;8(9):e1002894.

20. Antczak A, Kluźniak W, Wokołorczyk D, et al. A common nonsense mutation of the BLM gene and prostate cancer risk and survival. Gene 2013;532(2):173–176.

21. Baris HN, Kedar I, Halpern GJ, et al. Prevalence of breast and colorectal cancer in Ashkenazi Jewish carriers of Fanconi anemia and Bloom syndrome. Isr Med Assoc J 2007;9(12):847–850.

22. Cleary SP, Zhang W, Di Nicola N, et al. Heterozygosity for the BLM(Ash) mutation and cancer risk. Cancer Res 2003;63(8):1769–1771.

23. Kluźniak W, Wokołorczyk D, Rusak B, et al. Inherited Variants in BLM and the Risk and Clinical Characteristics of Breast Cancer. Cancers ;11(10.):

24. Gruber SB, Ellis NA, Scott KK, et al. BLM heterozygosity and the risk of colorectal cancer. Science 2002;297(5589):2013.

25. Bononi A, Goto K, Ak G, et al. Heterozygous germline *BLM* mutations increase susceptibility to asbestos and mesothelioma. Proc Natl Acad Sci U S A 2020;117(52):33466–33473.

26. Jaiswal S, Fontanillas P, Flannick J, et al. Age-related clonal hematopoiesis associated with adverse outcomes. N Engl J Med 2014;371(26):2488–2498.

27. Schenz J, Rump K, Siegler BH, et al. Increased prevalence of clonal hematopoiesis of indeterminate potential in hospitalized patients with COVID-19. Front Immunol 2022;13968778.

28. Steensma DP, Bejar R, Jaiswal S, et al. Clonal hematopoiesis of indeterminate potential and its distinction from myelodysplastic syndromes. Blood 2015;126(1):9–16.

29. Marnell CS, Bick A, Natarajan P. Clonal hematopoiesis of indeterminate potential (CHIP): Linking somatic mutations, hematopoiesis, chronic inflammation and cardiovascular disease. J Mol Cell Cardiol 2021;16198–105.

30. Yu B, Roberts MB, Raffield LM, et al. Supplemental Association of Clonal Hematopoiesis With Incident Heart Failure. J Am Coll Cardiol 2021;78(1):42–52.

31. Fuster JJ, Walsh K. Somatic Mutations and Clonal Hematopoiesis: Unexpected Potential New Drivers of Age-Related Cardiovascular Disease. Circ Res 2018;122(3):523–532.

32. Mitchell E, Spencer Chapman M, Williams N, et al. Clonal dynamics of haematopoiesis across the human lifespan. Nature 2022;606(7913):343–350.

33. Bick AG, Weinstock JS, Nandakumar SK, et al. Inherited causes of clonal haematopoiesis in 97,691 whole genomes. Nature 2020;586(7831):763–768.

34. Hagiwara K, Natarajan S, Wang Z, et al. Dynamics of age-versus therapy-related clonal hematopoiesis in long-term survivors of pediatric cancer. Cancer Discov [Epub ahead of print].

35. Loh P-R, Genovese G, Handsaker RE, et al. Insights into clonal haematopoiesis from 8,342 mosaic chromosomal alterations. Nature 2018;559(7714):350–355.

36. Loh P-R, Genovese G, McCarroll SA. Monogenic and polygenic inheritance become instruments for clonal selection. Nature 2020;584(7819):136–141.

37. DeBoy EA, Tassia MG, Schratz KE, et al. Familial Clonal Hematopoiesis in a Long Telomere Syndrome. N Engl J Med 2023;388(26):2422–2433.

38. Schratz KE, Haley L, Danoff SK, et al. Cancer spectrum and outcomes in the Mendelian short telomere syndromes. Blood 2020;135(22):1946–1956.

39. Schratz KE, Gaysinskaya V, Cosner ZL, et al. Somatic reversion impacts myelodysplastic syndromes and acute myeloid leukemia evolution in the short telomere disorders. J Clin Invest;131(18.):

40. Liu J, Tran D, Chan ICC, et al. Genetic Determinants of Clonal Hematopoiesis and Progression to Hematologic Malignancies in 479,117 Individuals.

41. Hoehn H, Salk D. Clonal analysis of stable chromosome rearrangements in Bloom’s syndrome fibroblasts. Cancer Genet Cytogenet 1984;11(4):405–415.

42. Vencio EF, Jenkins RB, Schiller JL, et al. Clonal cytogenetic abnormalities in Erdheim-Chester disease. Am J Surg Pathol 2007;31(2):319–321.

43. McKenna A, Hanna M, Banks E, et al. The Genome Analysis Toolkit: a MapReduce framework for analyzing next-generation DNA sequencing data. Genome Res 2010;20(9):1297–1303.

44. Nextera Flex for Enrichment Documentation. https://support.illumina.com/sequencing/sequencing_kits/nextera-flex-for-enrichment-\kit/documentation.html (accessed June 22, 2022).

45. Strom SP. Current practices and guidelines for clinical next-generation sequencing oncology testing. Cancer Biol Med 2016;13(1):3–11.

46. DeLeonardis K, Hogan L, Cannistra SA, Rangachari D, Tung N. When Should Tumor Genomic Profiling Prompt Consideration of Germline Testing? J Oncol Pract 2019;15(9):465–473.

47. Friedlaender A, Tsantoulis P, Chevallier M, De Vito C, Addeo A. The Impact of Variant Allele Frequency in EGFR Mutated NSCLC Patients on Targeted Therapy. Front Oncol 2021;11644472.

48. Coffee B, Cox HC, Bernhisel R, et al. A substantial proportion of apparently heterozygous TP53 pathogenic variants detected with a next-generation sequencing hereditary pan-cancer panel are acquired somatically. Hum Mutat 2020;41(1):203–211.

49. Wang Y, Bae T, Thorpe J, et al. Comprehensive identification of somatic nucleotide variants in human brain tissue. Genome Biol 2021;22(1):92.

50. Stacey SN, Zink F, Halldorsson GH, et al. Genetics and epidemiology of mutational barcode-defined clonal hematopoiesis. Nat Genet [Epub ahead of print].

51. Pan H, Renaud L, Chaligne R, et al. Discovery of Candidate DNA Methylation Cancer Driver Genes. Cancer Discov 2021;11(9):2266–2281.

52. Lee C-J, Ahn H, Jeong D, Pak M, Moon JH, Kim S. Impact of mutations in DNA methylation modification genes on genome-wide methylation landscapes and downstream gene activations in pan-cancer. BMC Med Genomics 2020;13(Suppl 3):27.

53. Levine ME, Lu AT, Quach A, et al. An epigenetic biomarker of aging for lifespan and healthspan. Aging 2018;10(4):573–591.

54. Lee J, Zhang J, Flanagan M, et al. Bloom syndrome patients and mice display accelerated epigenetic aging. Aging Cell 2023;e13964.

55. Guastafierro T, Bacalini MG, Marcoccia A, et al. Genome-wide DNA methylation analysis in blood cells from patients with Werner syndrome. Clin Epigenetics 2017;992.

56. Maierhofer A, Flunkert J, Oshima J, Martin GM, Haaf T, Horvath S. Accelerated epigenetic aging in Werner syndrome. Aging 2017;9(4):1143–1152.

57. Rasmussen KD, Helin K. Role of TET enzymes in DNA methylation, development, and cancer. Genes Dev 2016;30(7):733–750.

58. Ginno PA, Gaidatzis D, Feldmann A, et al. A genome-scale map of DNA methylation turnover identifies site-specific dependencies of DNMT and TET activity. Nat Commun 2020;11(1):2680.

59. Kong A, Frigge ML, Masson G, et al. Rate of de novo mutations and the importance of father’s age to disease risk. Nature 2012;488(7412):471–475.

60. Acuna-Hidalgo R, Sengul H, Steehouwer M, et al. Ultra-sensitive Sequencing Identifies High Prevalence of Clonal Hematopoiesis-Associated Mutations throughout Adult Life. Am J Hum Genet 2017;101(1):50–64.

61. Genovese G, Kähler AK, Handsaker RE, et al. Clonal hematopoiesis and blood-cancer risk inferred from blood DNA sequence. N Engl J Med 2014;371(26):2477–2487.

62. Conces M, Ni Y, Bazeley P, Patel B, Funchain P, Carraway HE. Clonal hematopoiesis of indeterminate potential (CHIP) mutations in solid tumor malignancies. J Clin Orthod 2019;37(15_suppl):1507–1507.

63. Climente-González H, Lonjou C, Lesueur F, et al. Boosting GWAS using biological networks: A study on susceptibility to familial breast cancer. PLoS Comput Biol 2021;17(3):e1008819.

64. Prokofyeva D, Bogdanova N, Dubrowinskaja N, et al. Nonsense mutation p.Q548X in BLM, the gene mutated in Bloom’s syndrome, is associated with breast cancer in Slavic populations. Breast Cancer Res Treat 2013;137(2):533–539.

65. Rashkin SR, Graff RE, Kachuri L, et al. Pan-cancer study detects genetic risk variants and shared genetic basis in two large cohorts. Nat Commun 2020;11(1):4423.

66. Laitman Y, Boker-Keinan L, Berkenstadt M, et al. The risk for developing cancer in Israeli ATM, BLM, and FANCC heterozygous mutation carriers. Cancer Genet 2016;209(3):70–74.

67. Hop PJ, Luijk R, Daxinger L, et al. Genome-wide identification of genes regulating DNA methylation using genetic anchors for causal inference. Genome Biol 2020;21(1):220.

68. Xie M, Lu C, Wang J, et al. Age-related mutations associated with clonal hematopoietic expansion and malignancies. Nat Med 2014;20(12):1472–1478.

69. Alhumaid M, Daher-Reyes GS, Schimmer AD, et al. Prognostic role of multiparameter flow cytometry-based measurable residual disease assessment in patients with acute myeloid leukemia harboring DNMT3A/TET2/ASXL1 mutation. Blood 2020;136(Supplement 1):8–9.

70. de Renty C, Ellis NA. Bloom’s syndrome: Why not premature aging?: A comparison of the BLM and WRN helicases. Ageing Res Rev 2017;3336–51.

71. Thomas M, Sukhai MA, Zhang T, et al. Integration of Technical, Bioinformatic, and Variant Assessment Approaches in the Validation of a Targeted Next-Generation Sequencing Panel for Myeloid Malignancies. Arch Pathol Lab Med 2017;141(6):759–775.

72. Sánchez R, Ayala R, Martínez-López J. Minimal Residual Disease Monitoring with Next-Generation Sequencing Methodologies in Hematological Malignancies. Int J Mol Sci;20(11.):

73. Li L, Eng C, Desnick RJ, German J, Ellis NA. Carrier frequency of the Bloom syndrome blmAsh mutation in the Ashkenazi Jewish population. Mol Genet Metab 1998;64(4):286– 290.

74. Shahrabani-Gargir L, Shomrat R, Yaron Y, Orr-Urtreger A, Groden J, Legum C. High frequency of a common Bloom syndrome Ashkenazi mutation among Jews of Polish origin. Genet Test 1998;2(4):293–296.

75. Levin MG, Nakao T, Zekavat SM, et al. Genetics of smoking and risk of clonal hematopoiesis. Sci Rep 2022;12(1):7248.

76. Bolton KL, Ptashkin RN, Gao T, et al. Cancer therapy shapes the fitness landscape of clonal hematopoiesis. Nat Genet 2020;52(11):1219–1226.

77. Armitage P, Doll R. The age distribution of cancer and a multi-stage theory of carcinogenesis. Br J Cancer 2004;91(12):1983–1989.

78. Goss KH, Risinger MA, Kordich JJ, et al. Enhanced tumor formation in mice heterozygous for Blm mutation. Science 2002;297(5589):2051–2053.

79. Hall MJ, Bernhisel R, Hughes E, et al. Germline Pathogenic Variants in the Ataxia Telangiectasia Mutated (ATM) Gene are Associated with High and Moderate Risks for Multiple Cancers. Cancer Prev Res 2021;14(4):433–440.

80. Del Valle J, Rofes P, Moreno-Cabrera JM, et al. Exploring the Role of Mutations in Fanconi Anemia Genes in Hereditary Cancer Patients. Cancers ;12(4.):

