## Supplemental Methods for "Increased Frequency of Clonal Hematopoiesis of Indeterminate Potential in Bloom Syndrome Probands and Carriers"

**Library Preparation.**

Genomic DNA was extracted from blood using standard Qiagen protocol. Exome sequencing was performed using the Nextera DNA Flex Pre-Enrichment Library Prep, followed by the Roche NimbleGen exome capture kit, according to established protocols. Libraries were indexed, multiplexed and sequenced on a 2x150 Illumina NovaSeq S1 flowcell at the UCLA Technology Center for Genomics and Pathology.

**Control Data.**

To assess the differences in *de novo* and somatic mutation rates, data from 19 control trio families were obtained through the database of Genotypes and Phenotypes (dbGaP). Control trios, with two parents and one child each, had unrelated conditions to cancer predisposition. Control children ranged from 0 to 17 years of age and control parents ranged from 19 to 60 years of age at time of collection (**Table S1**). Controls were phenotypically classified with metabolic disorders, mitochondrial disorders, or lung disease. Ten female children and nine male children control trios submitted from five institutions were randomly selected (**Table S1**).

**Bioinformatics Pipeline.**

All FASTQ files, with over 44 million unique reads and approximately 50% GC content, underwent stringent quality control^1^. The UCLA CDS DNA-alignment pipeline v6.1.0, which utilizes BWA-MEM2 (v2.1)^2^, was applied for alignments. GRCh38 assembly included additional contigs for Epstein-Barr virus (EBV), alternate haplotypes (ALT contigs), and human leukocyte antigen (HLA) loci for improved mapping and specificity^3^. Standard tools including BWA-MEM2 (v2.1)^2^, SAMtools (v1.10)^4^ and Picard Tools (v2.23.3)^5^ were used for alignment, sorting, duplicated marking and indexing. Nextera’s Flex for Enrichment exome kit was used to capture the targets.^6^ Base Quality Score Recalibration (BQSR), Depth of Coverage, Haplotype Caller, Variant Quality Score Recalibrator (VQSR), and contamination estimates were all performed using GATK (v4.2.4.1), while local realignments were performed using GATK (v3.7.0). The same versions of SAMtools and Picard were used to merge and reheader the BAMs at a chromosome level. Germline short variants (SNPs and Indels) were called using UCLA CDS germline SNP pipeline v5.4.3 which incorporated GATK4 best practices (**Figure S1A**).

**Variant Interpretation.**

Merged raw VCFs (variant call format) were processed using VarSeq v2.3.0 Exome Trio Template, filtering for read depth (DP)>10, genotype quality (GQ)>20 and VQSR = PASS. Variants were classified as de novo candidates, or transmitted variants based on standard VarSeq filters (**Figure S1B**).

**Coverage Validation.**

For CHIP and DNAm gene analysis, coverage was compared at DP>0, DP>10 and DP>50 across the four sample groups^7,8^. Loci for CHIP genes were visualized using a heatmap, ensuring consistency between exome capture kits.

**Variant Curation**.

Variants were sequentially filtered for 1) read depth (DP)>10 and genotype quality (GQ)>20 in the probands, 2) DP>10 and GQ>20 in parents; and 3) a quality score of “PASS” **(Figure S1B)**.

Exonic variants were retained, while intronic and intergenic variants were removed.

**Variant Allele Frequency (VAF) Analysis.**

VAF was calculated to distinguish between germline and somatic mutations. VAF<0.3 indicated acquired somatic variants, 0.3≤VAF≤0.7 represented heterozygous variants or *de novo* variants occurring germline, and 0.7<VAF≤1.0 represented homozygous reference alleles.

**Statistical analysis.**

Statistical analyses included normality checks, one-way (mixed) analysis of variance (ANOVA) model, where a random family effect was added to allow for non-independence between cohorts among members of the same family, for normally distributed data, non-parametric Kruskal-Wallis method for non-normally distributed data, and t-tests for comparing mean variant numbers. Multinomial logistic models with a random person effect were employed for analyzing mean proportions.Calculations were carried out using R, version 4.0.5.

**References**

1.	[Andrews. FastQC-A Quality Control application for FastQ files. Babraham Bioinformatics: Babraham, UK [Epub ahead of print].](http://paperpile.com/b/FlFlTK/Gs5Aq)

2.	Vasimuddin M, Misra S, Li H, Aluru S. Efficient Architecture-Aware Acceleration of BWA-MEM for Multicore Systems. *2019 IEEE International Parallel and Distributed Processing Symposium (IPDPS)*.

3.	GRCh38 - hg38 - Genome - Assembly - NCBI. <https://www.ncbi.nlm.nih.gov/assembly/GCF_000001405.26/> (accessed June 22, 2022).

4.	Li H, Handsaker B, Wysoker A, et al. The Sequence Alignment/Map format and SAMtools. Bioinformatics 2009;25(16):2078–2079.

5.	Picard. <http://broadinstitute.github.io/picard/> (accessed June 9, 2022).

6.	Nextera Flex for Enrichment Documentation. <https://support.illumina.com/sequencing/sequencing_kits/nextera-flex-for-enrichment-kit/documentation.html> (accessed June 22, 2022).

7.	Schenz J, Rump K, Siegler BH, et al. Increased prevalence of clonal hematopoiesis of indeterminate potential in hospitalized patients with COVID-19. Front Immunol 2022;13968778.

8.	[Hagiwara K, Natarajan S, Wang Z, et al. Dynamics of age- versus therapy-related clonal hematopoiesis in long-term survivors of pediatric cancer. Cancer Discov [Epub ahead of print].](http://paperpile.com/b/FlFlTK/hzI97)
