## Supplemental Figures for "Increased Frequency of Clonal Hematopoiesis of Indeterminate Potential in Bloom Syndrome Probands and Carriers"

**Supplemental Figures and Legends**

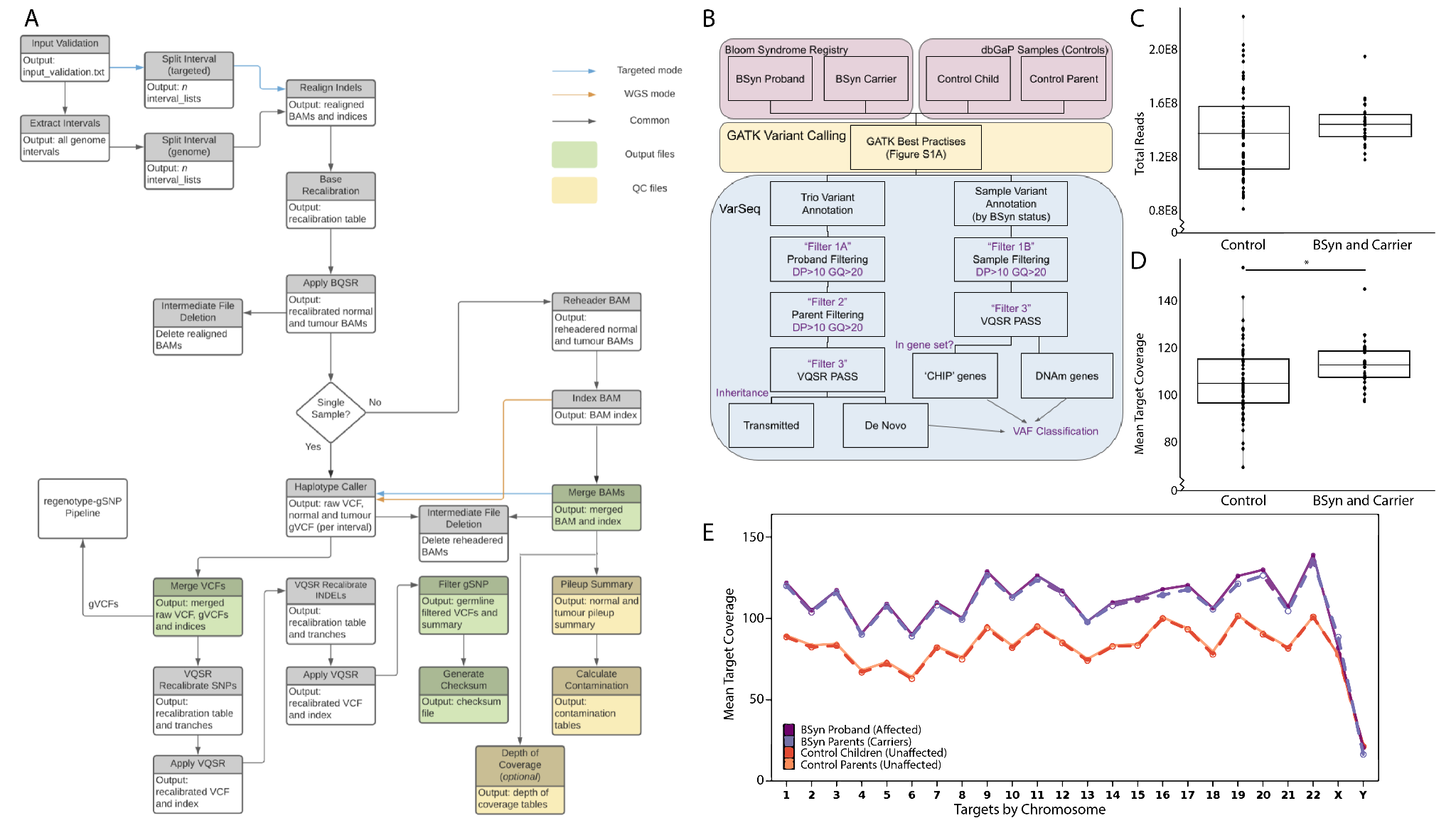

**Figure S1: Analysis pipelines and sample sequencing QC.**

(**A**) All exome files were processed using the UCLA CDS germline SNP pipeline v5.4.3 which incorporated GATK best practices. Base Quality Score Recalibration (BQSR) and Variant Quality Score Recalibration (VQSR) were performed using GATK v4.2.4.1 and local realignments were performed using GATK v3.7.0. (**B**) Merged raw VCFs generated from the UCLA CDS pipeline (yellow box) were then processed with the VarSeq v.2.3.0 Exome Trio Template (blue box). Annotated variants were then filtered for read depth (DP) and genotype quality (GQ) before classification (blue box). (**C**) We plotted total sequencing reads for each sample, and calculated mean total reads in the BSyn cohort (1.44 x 10^8^) and the control cohort (1.38 x 10^8^) (t-test, *p*-value=0.268). (**D**) We plotted exome coverage for each sample, and calculated mean exome coverage in BSyn cohort (113.1x) and the control cohort (106.5x) (t-test, *p*-value=0.018). (**E**) Mean exome coverage between the cohorts - BSyn proband (n=10), and carrier (n=19), control children (n=19), and parents (n=38) - at each chromosome was plotted to examine any chromosome-specific enrichment in exome coverage.

*p*-values were determined by t-test, and ns denote *p*-value>.05, * denote *p*-value≤.05

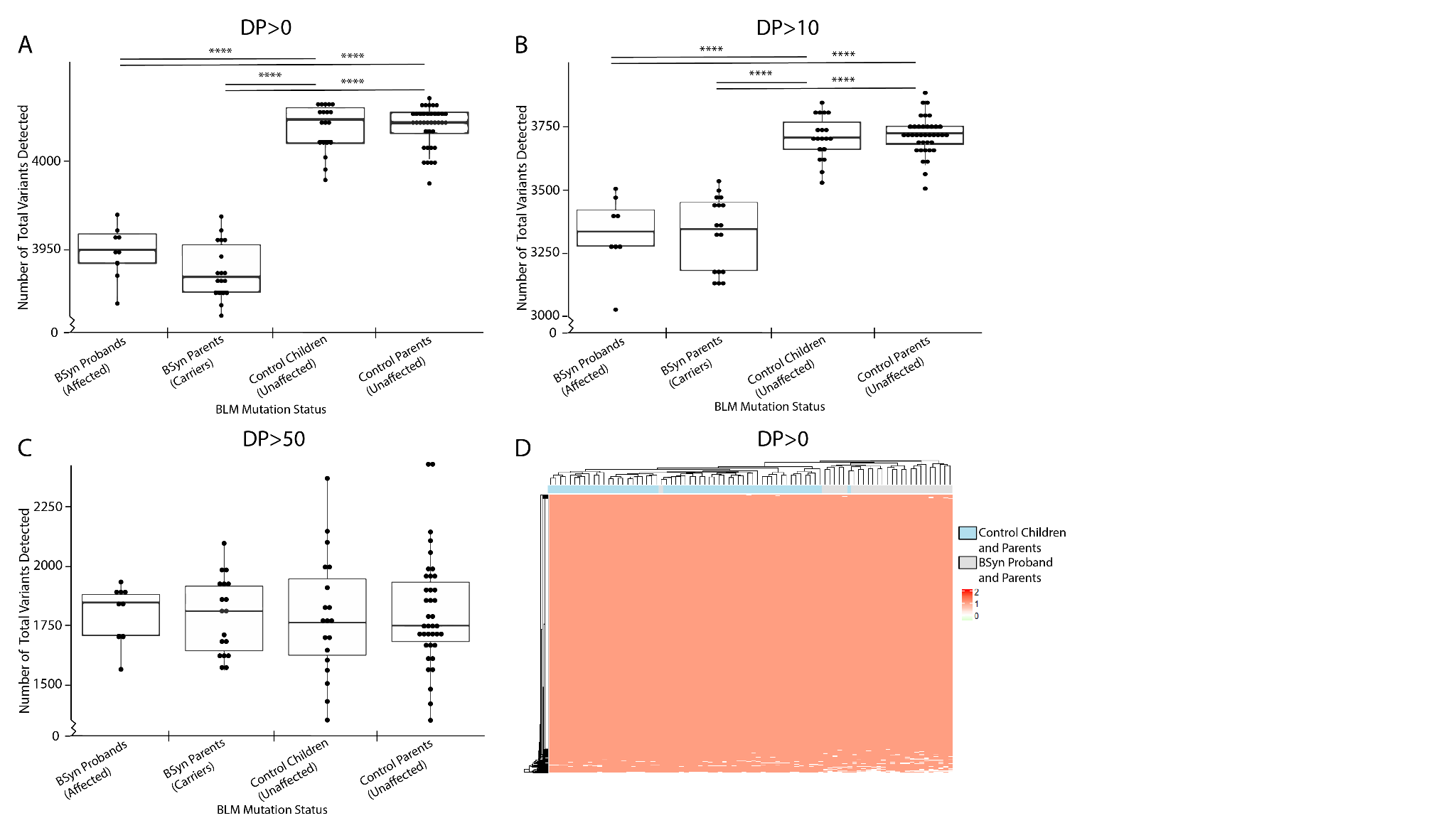

**Figure S2: Coverage QC for CHIP Genes**

The total number of variants detected for each sample in the subset of CHIP genes was identified at a read depth of (**A**) DP>0 (t-test, significant *p*-values range from 6.84E-21 to 2.65E-34), (**B**) DP>10 (t-test, significant *p*-values range from 2.11E-12 to 4.30 E-20) and (**C**) DP>50 (t-test, no significant *p*-values) and compared between the four sample groups. (**D**) Each loci covered at DP>0 was plotted as a heat map across all samples to compare capture.

*p*-values were determined by t-test, ns or no stars denote *p*-value>.05, * denote *p*-value≤.05, ** denote *p*-value<.01, *** denote *p*-value<.001, **** denote *p*-value<.0001

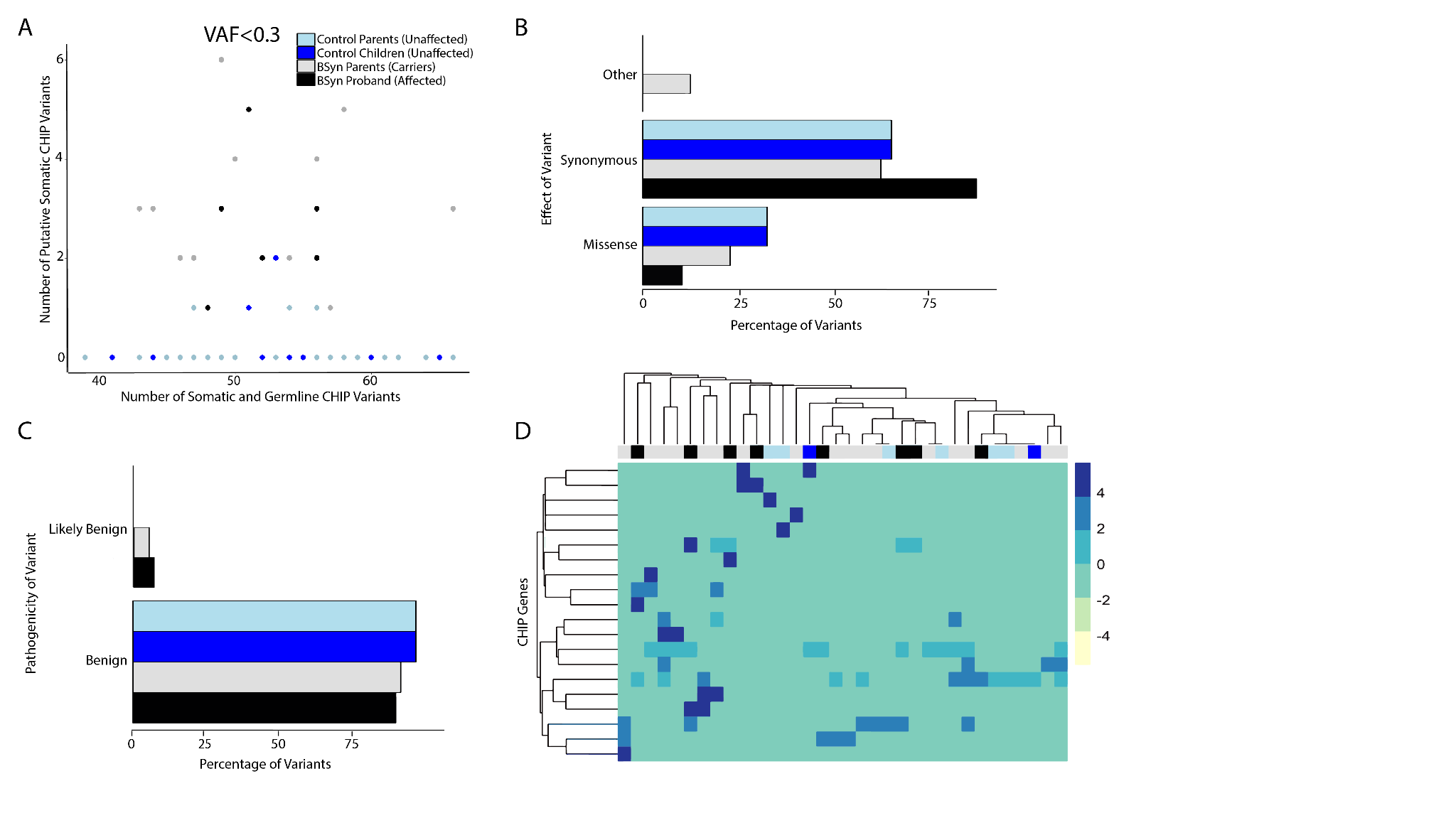

**Figure S3: Increased CHIP variant load in BSyn probands and carriers does not appear to cause deleterious genomic effects.**

(**A**) Total number of CHIP gene variants identified in a sample compared to the number of somatic CHIP gene variants. (**B**) We examined the consequences (RefSeq Genes 110, NCBI) of the putative somatic CHIP variants, specifically synonymous, missense, loss of function (LoF) or other (splice etc.) and (**C**) analyzed breakdown based on pathogenicity (ClinVar 2023-01-05, NCBI). (**D**) We analyzed breakdown of putative somatic CHIP variants across all 56 CHIP genes (Table S5, RefSeq Genes 110, NCBI) identified in literature, organized based on gene of variation (y axis) and by sample (x axis) using hierarchical clustering. The heatmap depicts the distribution of putative somatic CHIP variants at VAF < 0.3 identified in the samples, with each row representing a CHIP gene and each column representing a sample. Despite the hierarchical clustering, there are no discernible relationships between the sample cohort and the genes where variants were identified, suggesting a heterogeneous pattern of putative somatic CHIP gene variants across the samples.

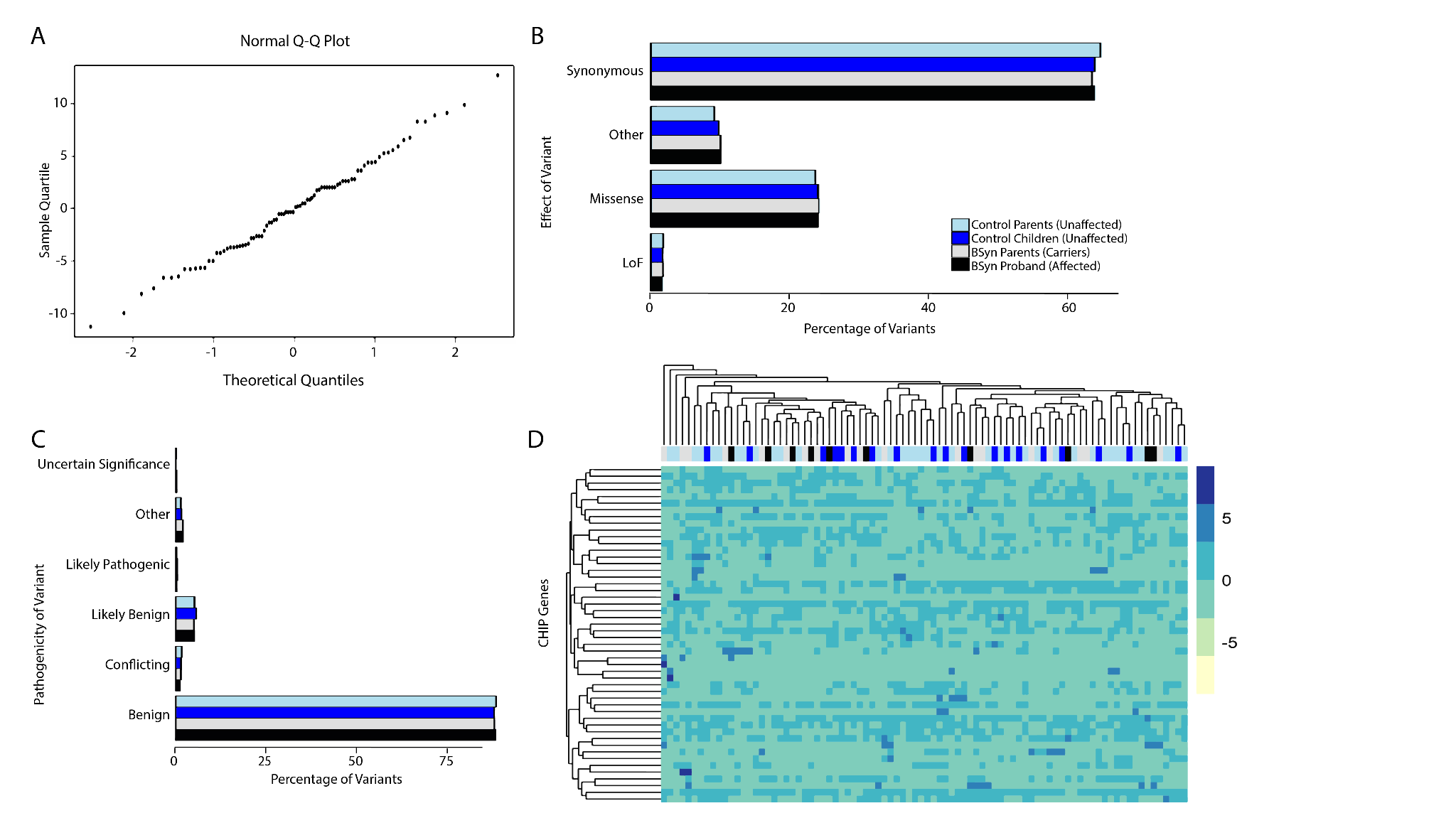

**Figure S4: Total variants in CHIP genes in BSyn and control cohorts are normally distributed.**

(**A**) A Quantile-Quantile (Q-Q) plot is presented to assess the normal distribution of total CHIP gene variants for each sample. The straight line observed in the plot indicates that the distribution of variants closely follows a normal distribution, suggesting that the assumption of normality is valid for the distribution of CHIP gene variants across all samples. Each point on the plot represents a quantile of the observed data compared to the expected quantile of a normal distribution. (**B**) We examined the consequences (RefSeq Genes 110, NCBI) of the CHIP gene variants, specifically synonymous, missense, loss of function (LoF) or other (splice etc.) and (**C**) analyzed breakdown based on pathogenicity (ClinVar 2023-01-05, NCBI). (**D**) We analyzed breakdown of CHIP gene variants across all 56 CHIP genes (Table S5, RefSeq Genes 110, NCBI) identified in literature based on gene of variation (y axis) and by sample (x axis) using hierarchical clustering. The heatmap depicts the distribution of all CHIP variants regardless of VAF identified in the samples, with each row representing a CHIP gene and each column representing a sample. Despite the hierarchical clustering, there are no discernible relationships between the sample cohort and the genes where variants were identified, suggesting a heterogeneous pattern of CHIP gene variants across the samples.

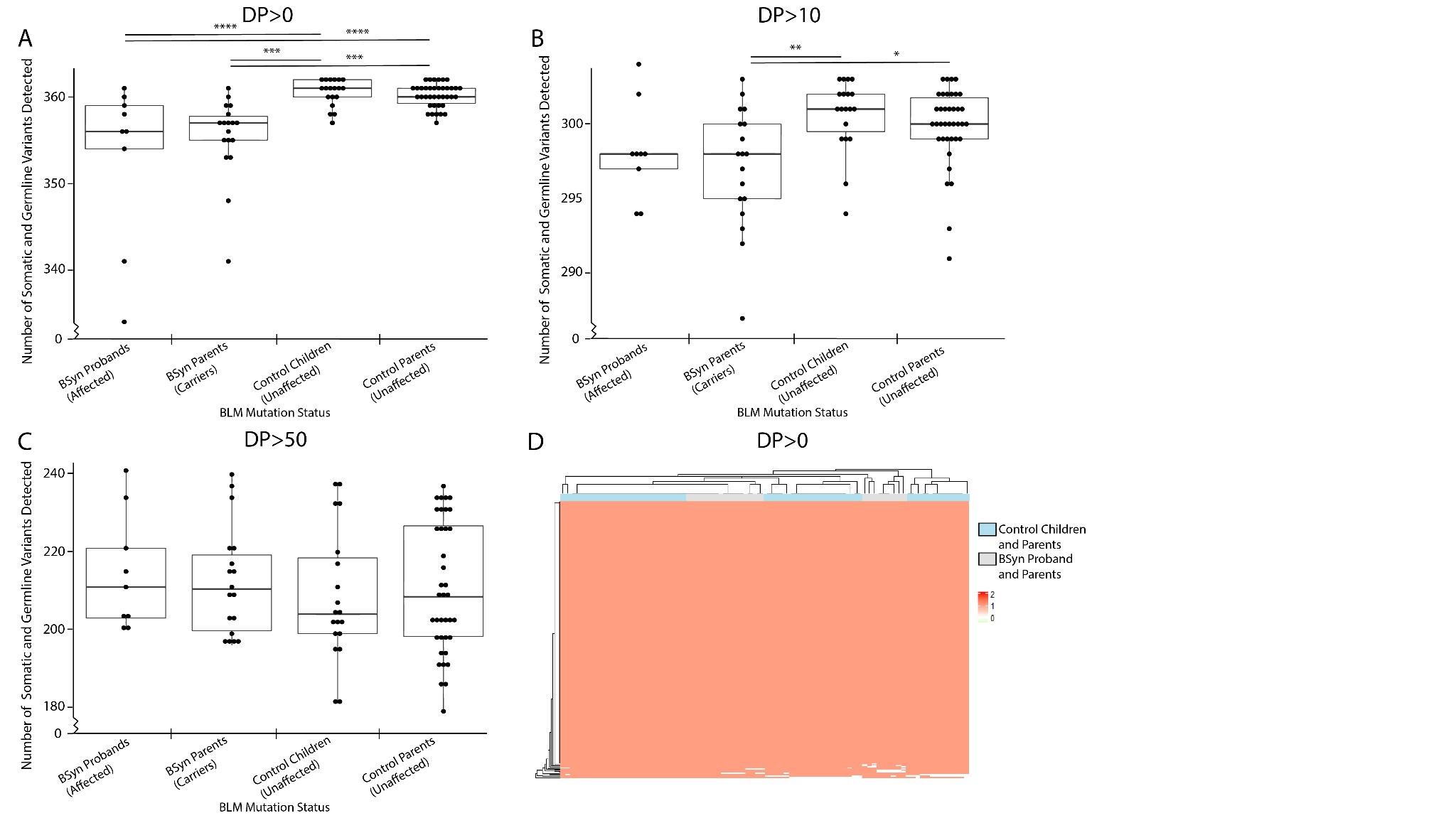

**Figure S5: Coverage QC for DNAm Genes**

The total number of variants detected for each sample in the subset of DNAm genes was identified at a read depth of (**A**) DP>0 (t-test, significant *p*-values range from 1.30E-04 to 5.13E-06), (**B**) DP>10 (t-test, significant *p*-values range from 0.012 to 0.004) and (**C**) DP>50 (t-test, no significant *p*-values) and compared between the four sample groups. (**D**) Each loci covered at DP>0 was plotted as a heat map across all samples to compare capture.

*p*-values were determined by t-test, ns or no stars denote *p*-value>.05, * denote *p*-value≤.05, ** denote *p*-value<.01, *** denote *p*-value<.001, **** denote *p*-value<.0001

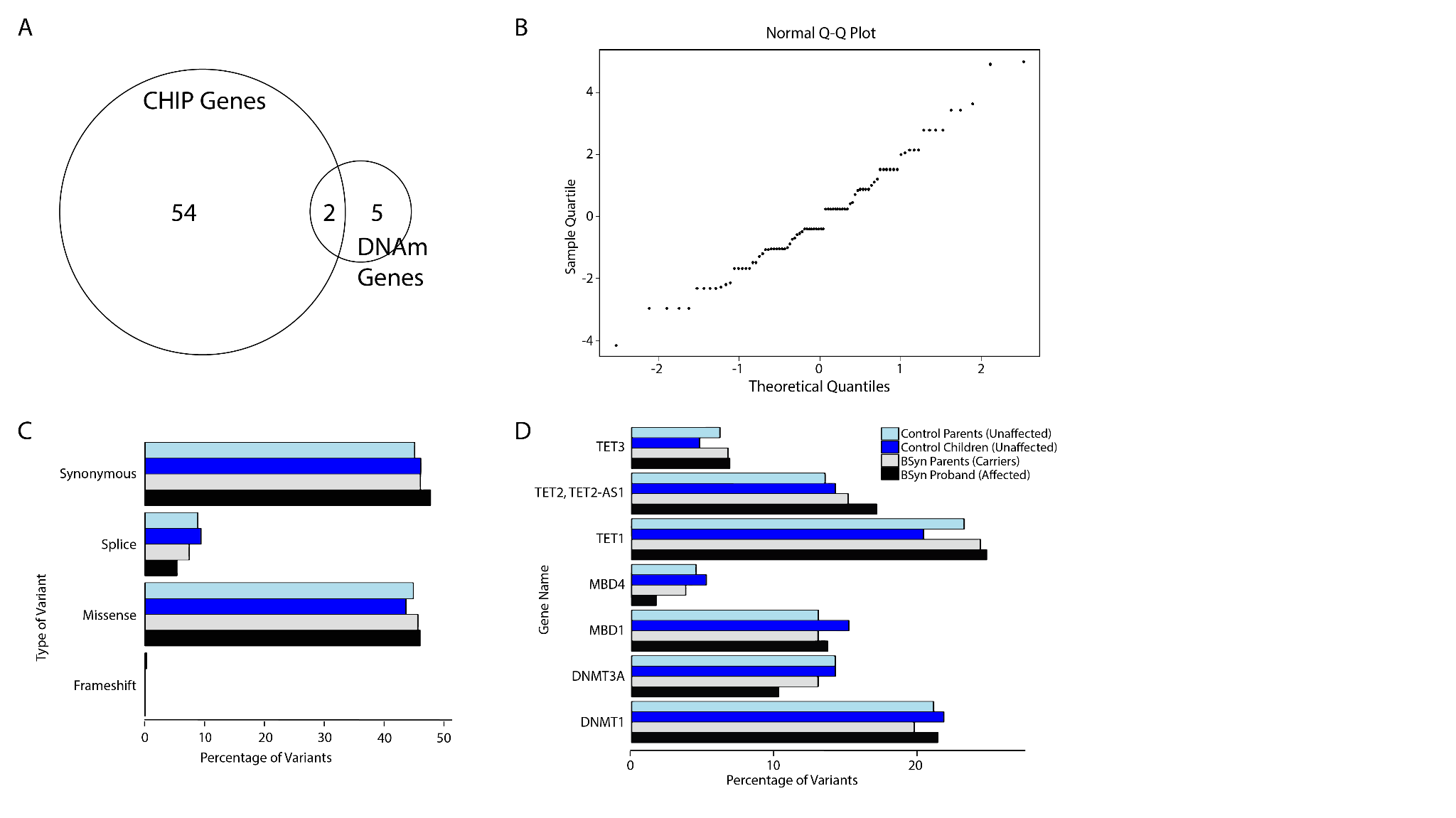

**Figure S6: Total variants in DNAm genes in BSyn and control cohorts are normally distributed.**

(**A**) Genes involved in DNA methylation (DNAm) or implicated in CHIP (Table S5) have slight overlap. (**B**) A Quantile-Quantile (Q-Q) plot is shown to evaluate the normal distribution of total DNAm gene variants for each sample. The straight line in the plot signifies that the distribution of variants adheres to a normal distribution. The points on the plot represent quantiles of the observed data in comparison to the expected quantiles of a normal distribution. (**C**) We examined the consequences (RefSeq Genes 110, NCBI) of the DNAm gene variants, specifically synonymous, missense, loss of function (LoF) or other (splice etc.). (**D**) We analyzed breakdown of DNAm gene variants across all 7 DNAm genes (Table S5, RefSeq Genes 110, NCBI) identified in literature based on gene of variation (y axis) and by sample (x axis) using unsupervised clustering.

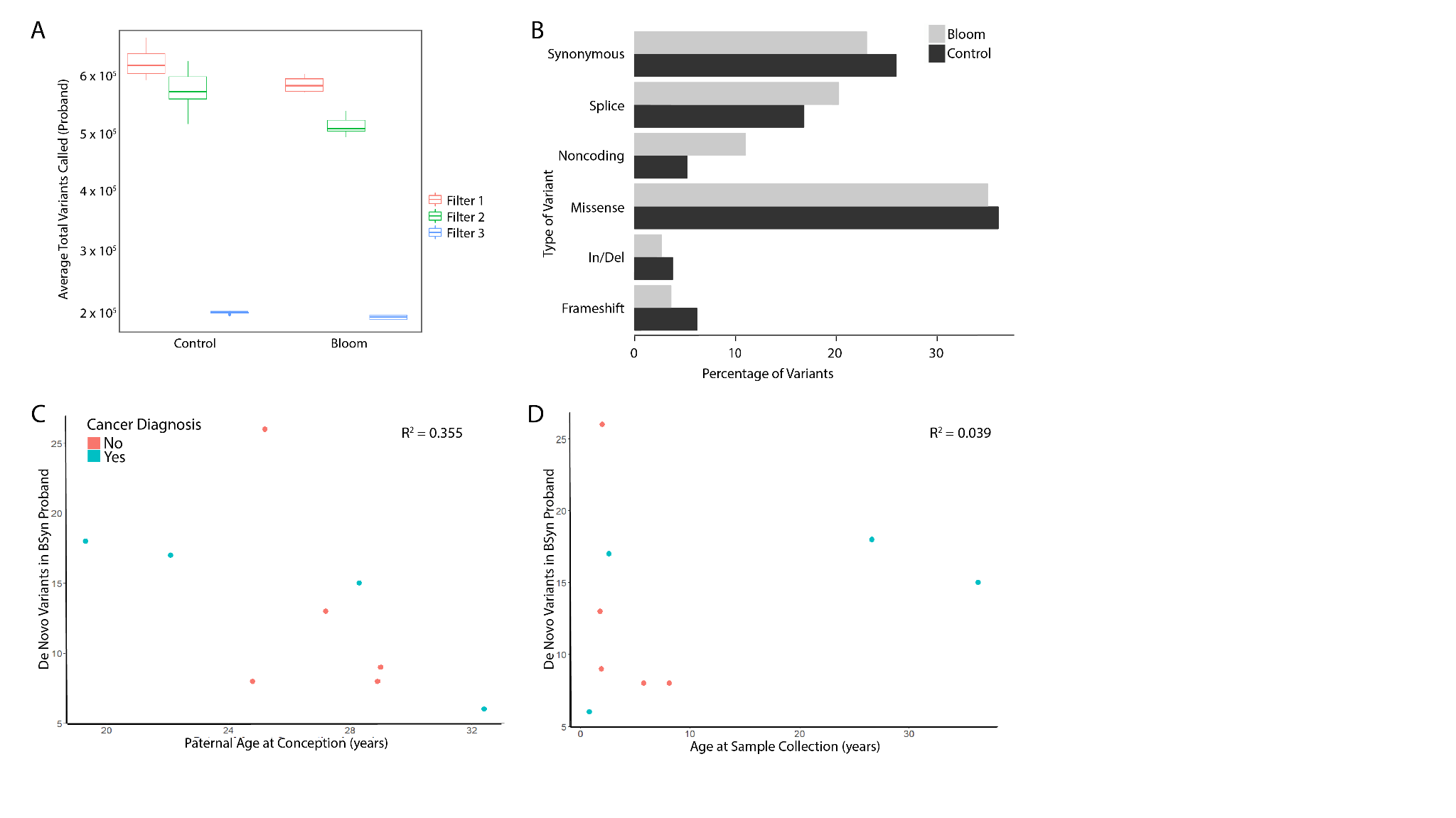

**Figure S7: Trio exome analysis did not identify significant trends in *de novo* variants in BSyn probands.**

(**A**) Trio exome samples for BSyn probands (n=9) and control children (n=19) were annotated and processed in VarSeq v.2.3.0 using read depth, genotype quality, and variant PASS filters (see Methods). (**B**) We examined the consequences (RefSeq Genes 110, NCBI) of the *de novo* variants, such as synonymous, missense, loss of function (LoF) or other (splice etc.). (**C**) Linear regression analysis was performed to assess the effect of paternal age at conception on rate of *de novo* variants in the BSyn probands (R^2^=0.355), (**D**) and the effect of BSyn proband age at sample collection on number of *de novo* variants (R^2^=0.039).
